# Using deep learning algorithms to simultaneously identify right and left ventricular dysfunction from the electrocardiogram

**DOI:** 10.1101/2021.03.31.21254696

**Authors:** Akhil Vaid, Kipp W Johnson, Marcus A Badgeley, Sulaiman S Somani, Mesude Bicak, Isotta Landi, Adam Russak, Shan Zhao, Matthew Levin, Robert S Freeman, Alexander W Charney, Atul Kukar, Bette Kim, Tatyana Danilov, Stamatios Lerakis, Edgar Argulian, Jagat Narula, Girish N Nadkarni, Benjamin S Glicksberg

## Abstract

**Background:** Rapid evaluation of left and right ventricular function using deep learning (DL) on electrocardiograms (ECG) can assist diagnostic workflow. However, DL tools to estimate right ventricular (RV) function do not exist, while ones to estimate left ventricular (LV) function are restricted to quantification of very low LV function only.

**Objectives:** This study sought to develop deep learning models capable of comprehensively quantifying left and right ventricular dysfunction from ECG data in a large, diverse population.

**Methods:** A multi-center study was conducted with data from five New York City hospitals; four for internal testing and one serving as external validation. We created novel DL models to classify Left Ventricular Ejection Fraction (LVEF) into categories derived from the latest universal definition of heart failure, estimate LVEF through regression, and predict a composite outcome of either RV systolic dysfunction or RV dilation.

**Results:** We obtained echocardiogram LVEF estimates for 147,636 patients paired to 715,890 ECGs. We used Natural Language Processing (NLP) to extract RV size and systolic function information from 404,502 echocardiogram reports paired to 761,510 ECGs for 148,227 patients.

For LVEF classification in internal testing, Area Under Curve (AUC) at detection of LVEF<=40%, 40%<LVEF<=50%, and LVEF>50% was 0.94 (95% CI:0.94-0.94), 0.82 (0.81-0.83), and 0.89 (0.89-0.89) respectively. For external validation, these results were 0.94 (0.94-0.95), 0.73 (0.72-0.74) and 0.87 (0.87-0.88). For regression, the mean absolute error was 5.84% (5.82-5.85) for internal testing, and 6.14% (6.13-6.16) in external validation. For prediction of the composite RV outcome, AUC was 0.84 (0.84-0.84) in both internal testing and external validation.

**Conclusions:** DL on ECG data can be utilized to create inexpensive screening, diagnostic, and predictive tools for both LV/RV dysfunction. Such tools may bridge the applicability of ECGs and echocardiography, and enable prioritization of patients for further interventions for either sided failure progressing to biventricular disease.

## Introduction

Heart failure represents a significant health burden, with an estimated 6.2 million people affected in the United States^1^, and at least 64 million people worldwide^2^. Considerable attention has been paid to the pathophysiology of left ventricular (LV) failure. However, owing to the anatomical and functional proximity of the ventricles, either LV or right ventricular (RV) failure can precipitate biventricular involvement, with even subclinical RV dysfunction having been found to be associated with the risk of LV failure^3^. Patients with biventricular failure have significantly worse outcomes, with 2-year survival of 23%, as opposed to 71% in patients with isolated LV failure^3,4^. Studies also show that RV dysfunction is indicative of prognosis independent of LV dysfunction for a plethora of cardiac diseases^5-16^. In recognition of the inextricable nature of LV disease, RV disease, and biventricular disease, the recent first Universal Definition and Classification of Heart Failure did not attempt to specify distinct clinical entities for left and right heart failure^17^.

Early detection of heart failure creates the possibility of more efficient implementation of guideline-directed medical therapy and lifestyle modifications, which have been shown to improve overall outcomes for all forms of heart failure, in addition to slowing progression to advanced disease.^18^

Left ventricular ejection fraction (LVEF) is one of the most widespread hemodynamic parameters currently available in cardiovascular medicine. Among many other possible uses, LVEF as a measure of ventricular function, can be used to quantify disease progression and response to treatment^19-21^, and to independently predict mortality^22^. LVEF measurements are most readily obtained by transthoracic echocardiography, and thus echocardiography is one of the procedures most commonly billed to Medicare and Medicaid in the United States^23^. However, significant barriers remain to obtaining LVEF measurements in outpatient or resource-limited settings without sufficient trained echocardiographers and logistical support^24^, and there remains significant inter-observer and intra-observer variability in measurement^17^. Furthermore, trajectories of LVEF over time may be more useful than isolated measurements^17^ requiring repeated echocardiograms and trips to the clinic for patients.

In contrast, RV failure has classically been within the realm of clinical diagnosis, with no specific biomarkers or agreed upon guidelines for ECG interpretation. Numerical measurements of RV function such as RV ejection fraction are not as readily available due to difficulties in measurement from conventional transthoracic echocardiography^25^. Alternate methods to assess RV function such as tricuspid annular systolic plane excursion (TAPSE) have demonstrated promise in certain clinical settings^26^, but there remain challenges in common scenarios such as measuring disease progression^27^ or assessment of RV function following cardiac surgery^28^. 3D echocardiography, strain imaging, and cardiac MRI are promising replacements^29^, but are impractical for use as screening modalities due to concerns of cost and availability. Thus, assessment of the important role of right ventricular function in the pathophysiology of cardiovascular disease has to date been underappreciated^30^.

Taken together, there exists a pressing need for a readily available and inexpensive tool to simultaneously measure, screen, or predict both right and left heart function. The ECG is a cardinal investigation in the practice of Cardiology. It is ubiquitous, inexpensive and is often the first investigation performed in emergency situations. However, it has an upper bound of usefulness secondary to its skill requirement and subjectivity. Additionally, clinicians cannot identify subtle patterns that might indicate sub-clinical pathology, especially for conditions for which there are no interpretation guidelines. Recent breakthroughs in artificial intelligence have demonstrated that much more information may be available from the ECG to diagnose such conditions than currently leveraged^31,32^. Deep learning (DL), a class of machine learning that uses hierarchical networks to extract lower-dimensional features from a higher dimensional data input, has demonstrated significant potential for enabling ECG-based predictions and diagnoses^33^. For example, DL has been used to identify patients with atrial fibrillation while in normal sinus rhythm^34^, predict incident atrial fibrillation^35^, identify patients amenable to cardiac resynchronization therapy^36^, evaluate LV diastolic function^37^, evaluation of patients with echocardiographically concealed long QT syndrome^38^, predict risk of sudden cardiac death^39^, and to predict low LVEF.^40,41^.

For the first time, we present novel DL algorithms using ECG waveform data to simultaneously predict the presence of both left ventricular and right ventricular disease in a large, ethnically and socioeconomically diverse population. We further establish a regression framework to predict numerical values of LVEF. Finally, we evaluate these algorithms with respect to mitral valve regurgitation and in clinically meaningful ranges of LVEF.

## Methods

### Data Source and Patient Population

We utilized patient data from five hospitals within the Mount Sinai Health System from 2003-2020. These hospitals, specifically Mount Sinai Hospital, Mount Sinai Morningside, Mount Sinai Brooklyn, Mont Sinai West, and Mount Sinai Beth Israel, are located in Manhattan, and Brooklyn boroughs of New York City and serve a large and diverse patient population. **Table 1** reflects the patient population utilized in this work. **Figure 1** displays the workflow of obtaining and processing relevant data. We extracted Left Ventricular Ejection Fraction (LVEF) from the EPIC electronic healthcare record (EHR) system. LVEF values are abstracted from transthoracic echocardiography (echo) result reports written by physicians. Extracted records contained a unique identifier for the patient, the date and time of the echo, and the value of the LVEF as an integer. Utilizing this method, we acquired 444,786 reports for 219,437 patients.

**Table 1:** Population metrics of the study cohort by Left Ventricular Ejection Fraction classification

|  | LVEF ≤ 40% | 40% < LVEF ≤ 50% | LVEF > 50% | p |
| --- | --- | --- | --- | --- |
| **Internal Testing Cohort** |  |  |  |  |
| Patients: n | 17,840 | 19,586 | 109,615 |  |
| Echo-ECG pairs: n | 171,017 | 112,300 | 431,134 |  |
| Age | 67.0 (66.9 - 67.1) | 66.6 (66.5 - 66.6) | 65.6 (65.6 - 65.7) | <0.0001 |
| Gender n (%) |  |  |  | <0.0001 |
| Male | 12,230 (68.55) | 13,128 (67.03) | 55,831 (50.93) |  |
| Female | 5,610 (31.45) | 6,458 (32.97) | 53,784 (49.07) |  |
| Race n (%) |  |  |  | <0.0001 |
| American Indian | 419 (2.35%) | 552 (2.82%) | 2,755 (2.51%) |  |
| Asian | 505 (2.83%) | 556 (2.84%) | 3,962 (3.61%) |  |
| Black | 1,865 (10.45%) | 1,883 (9.61%) | 9,025 (8.23%) |  |
| Hispanic | 1,975 (11.07%) | 2,196 (11.21%) | 11,300 (10.31%) |  |
| Other | 1,302 (7.3%) | 1,365 (6.97%) | 8,041 (7.34%) |  |
| Pacific Islander | 41 (0.23%) | 24 (0.12%) | 126 (0.11%) |  |
| Unknown | 7,092 (39.75%) | 7,669 (39.16%) | 48,591 (44.33%) |  |
| White | 4,641 (26.01%) | 5,341 (27.27%) | 25,815 (23.55%) |  |
| LVEF | 28.7 (28.6 - 28.7) | 46.8 (46.7 - 46.8) | 62.0 (62.0 - 62.0) | <0.0001 |
| Ventricular Rate | 80.8 (80.5 - 81.0) | 76.3 (76.1 - 76.6) | 74.2 (74.1 - 74.3) | <0.0001 |
| Atrial Rate | 92.0 (91.2 - 92.9) | 85.5 (84.8 - 86.2) | 79.3 (79.1 - 79.5) | <0.0001 |
| PR Interval | 170.6 (167.7 - 173.5) | 168.6 (168.0 - 169.1) | 163.7 (163.5 - 163.9) | <0.0001 |
| QTc Interval | 471.0 (470.3 - 471.6) | 453.8 (453.2 - 454.3) | 439.4 (439.2 - 439.6) | <0.0001 |
| **External Validation Cohort** |  |  |  |  |
| Patients | 123 | 84 | 390 |  |
| Echo-ECG pairs | 372 | 214 | 853 |  |
| Age | 64.0 (62.4 - 65.6) | 63.8 (61.8 - 65.8) | 62.3 (61.3 - 63.4) | 0.15 |
| Gender n (%) |  |  |  | <0.0001 |
| Male | 81 (65.85) | 58 (69.05) | 200 (51.28%) |  |
| Female | 42 (34.15) | 26 (30.95) | 190 (48.72%) |  |
| Race n (%) |  |  |  | <0.0001 |
| American Indian | - | - | - |  |
| Asian | - | - | - |  |
| Black | 14 (11.38%) | 12 (14.29%) | 81 (23.21%) |  |
| Hispanic | - | - | 25 (7.16%) |  |
| Other | 32 (26.02%) | 20 (23.81%) | 90 (25.79%) |  |
| Pacific Islander | - | - | - |  |
| Unknown | 38 (30.89%) | 21 (25.0%) | 97 (24.87%) |  |
| White | 28 (22.76%) | 24 (28.57%) | 95 (27.22%) |  |
| LVEF | 25.5 (24.6 - 26.3) | 47.3 (46.9 - 47.7) | 61.5 (61.1 - 61.8) | <0.0001 |
| Ventricular Rate | 84.4 (80.9 - 87.9) | 78.5 (75.4 - 81.5) | 79.8 (78.1 - 81.6) | <0.0001 |
| Atrial Rate | 90.9 (83.1 - 98.8) | 93.7 (79.0 - 108.5) | 86.1 (82.1 - 90.1) | <0.0001 |
| PR Interval | 167.8 (160.4 - 175.1) | 175.6 (165.7 - 185.5) | 161.4 (158.2 - 164.5) | <0.0001 |
| QTc Interval | 480.9 (473.0 - 488.8) | 465.0 (456.9 - 473.2) | 448.6 (444.8 - 452.3) | <0.0001 |

**Figure 1:**
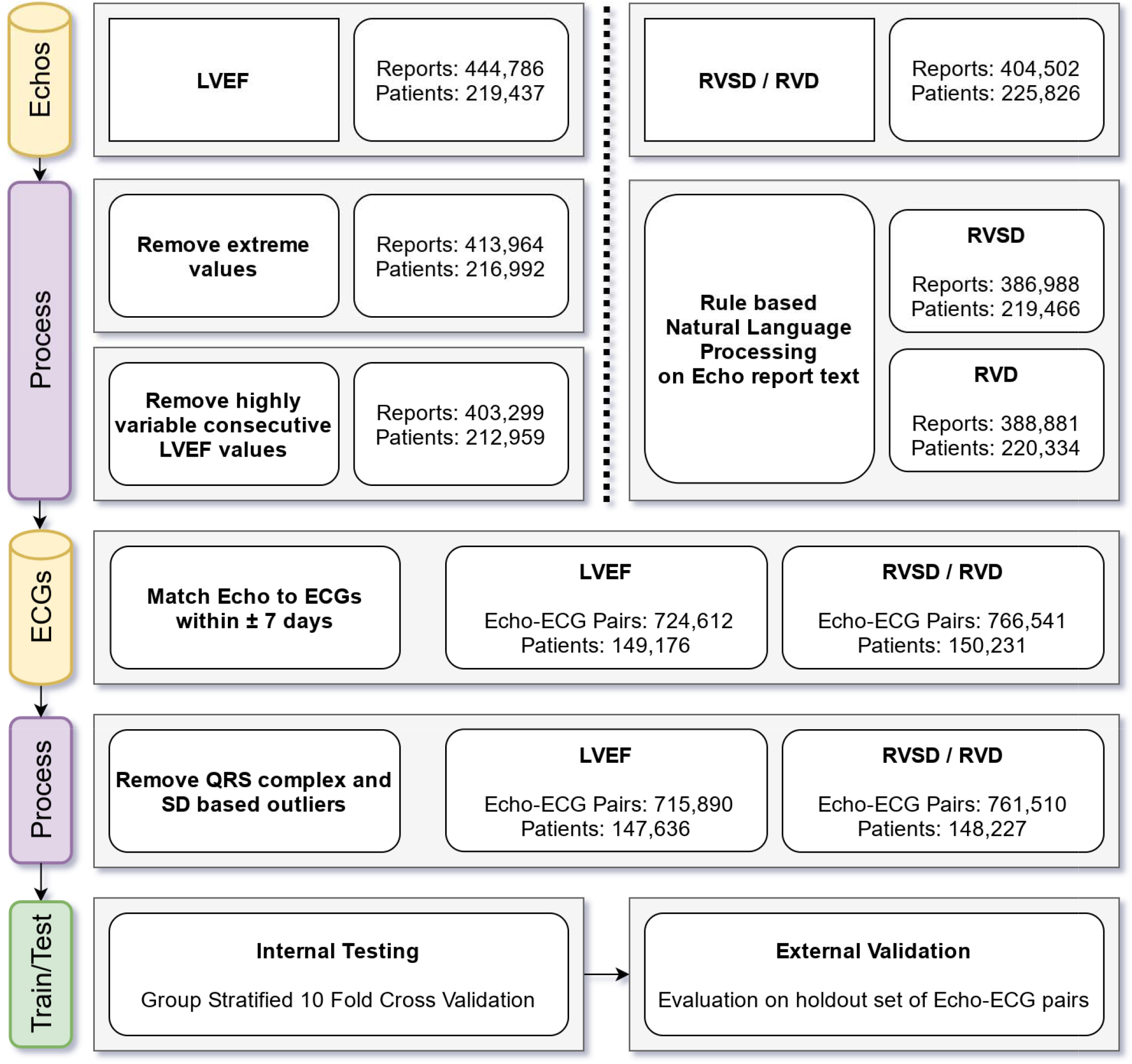
CONSORT Diagram. Sources of data are highlighted in yellow. Data processing steps are highlighted in purple. Numbers in each step indicate datapoints retained for analysis following conditional filtering. *LVEF*: Left Ventricular Ejection Fraction, *RVSD*: Right Ventricular Systolic Dysfunction, *RVD*: Right Ventricular Dilation, *SD*: Standard Deviation.

Details of either Right Ventricular Systolic Dysfunction (RVSD) or Right Ventricular Dilation (RVD) were not present as discrete parameters within the EHR database. We acquired .pdf files containing unstructured text corresponding to 404,502 transthoracic echo reports for 225,826 patients. As before, collected records also contained a unique identifier for the patient, and the date of the echo.

ECG data were obtained as XML files (*eXtensible Markup Language*) exported from the GE MUSE ECG system. These files contain demographic details for the patient, details about the testing site, per-lead cart generated parameters for ECGs, cart generated ECG diagnosis, and raw waveform data (see *Electrocardiograph* data subsection for more details). For each outcome defined by an echo as described below, we paired the echo report to any ECG performed within a time period of 7 days before, to 7 days after the date of the echo. Overall, we extracted 715,890 paired ECGs for 147,636 patients for LVEF prediction, and 761,510 paired ECGs for 148,227 patients for Right Ventricular status prediction. Finally, there was an overlap of 390,921 paired ECGs for 87,514 patients over the two datasets. **(Figure 1)**

### Definition of Primary Outcomes

First, we elected to model LVEF in a classification framework. LVEF was stratified into three clinically relevant ranges of LVEF <= 40%, LVEF > 40% and <= 50%, and LVEF > 50%.^42^ Since none of these intervals overlap, the overall task may be considered a multi-class classification problem. To compare to prior work, we also assessed performance at classification of LVEF <= 35%. Second, we attempted to model LVEF using a regression framework (i.e., directly predicting integer values of LVEF). For this problem, the target label was the LVEF value associated with each Echo-ECG pair and required no additional processing.

Right heart status was considered as a composite phenotype positive for either Right Ventricular Systolic Dysfunction (RVSD) or Right Ventricular Dilation (RVD) as elicited from an echo report. The process for defining right heart status relied on use of natural language processing (NLP) of the text from the report (*see Natural Language Processing of Echocardiography Reports for Outcome Extraction*). Phrases used to define either of RVSD and RVD are listed in **Supplementary Table 1**. An Echo-ECG pair was labelled positive for the outcome and assigned a value of 1 in the presence of either RVD or RVSD of any severity, or a value of 0 if both were absent. Since there were only two possible values for the outcome, the task may be considered a binary classification problem.

### Data Processing, Quality Control, and Filtering

#### Left Ventricular Ejection Fraction

We discarded outliers above 90% LVEF (99.77^th^ percentile), and below 10% LVEF (0.18^th^ percentile) within our patient population. **(Figure 1: CONSORT diagram)** Additionally, the value of LVEF generated from echo is subject to inter-rater and inter-test variability. Since we considered data collected over a period of ±7 days, if the difference in reported LVEF for a patient between any two consecutive reports within 7 days was greater than 10%^43^, both of these reports were discarded.

#### Natural Language Processing of Echocardiography Reports for Outcome Extraction

We chose a rule-based approach for extracting outcomes of interest from the text contained within echo reports - specifically Right Ventricular Systolic Dysfunction (RVSD), Right Ventricular Dilation (RVD), and Mitral Regurgitation. We created and iteratively expanded an overall list of rules designed to ensure capture of the variability surrounding phrases detailing the same semantic concept. We provide the final list of rules in **Supplementary Table 1**, and anonymized sample annotated echo reports in **Supplementary Figures 1, 2, and 3**.

While RVD and RVSD were only considered in terms of their presence or absence, we created additional rules to extract qualifiers of valvular disease severity. A total of 8 rules were created to be able to classify Mitral Regurgitation into Normal, Borderline (Trace/Minimal/Mild), Moderate, or Severe disease.

Overall NLP performance was measured through two single-blind faculty reviewers. Each review contained 210 reports randomly sampled and equally distributed based on detected normal, detected diseased for either of RVD, RVSD or MR, and no mention detected for RV or Mitral Valve status.

#### Electrocardiography Data

Waveform data within XML files is formatted as one-dimensional collections (vectors) of integers sampled at a rate of 500Hz. Each such vector corresponds to a lead, with each XML file containing data for leads I, II, and V1 - V6. These vectors extend to either 5 seconds (2500 samples), or 10 seconds (5000 samples) of recorded information for each lead, in addition to longer rhythm strip recordings. To avoid potential artifacts caused by extending 2500 samples to 5000, we restricted each sample to only the first 5 seconds of recording. Furthermore, our dataset does not contain data regarding leads III, aVF, aVL, or aVR. For our purposes, these leads were considered to have no additional information since they can be derived from linear transformations of the vectors representing the other leads^44^, and were therefore not included in our models.

ECGs are prone to recording errors such as baseline wander and electrical interference^45^. We corrected for these through the application of a median filter applied over a 2 second window, and a subsequent Butterworth Bandpass filter applied to a 0.5 - 40 Hz range. In order to account for recordings with no lead information, or excess noise despite filtering, we calculated the average QRS amplitude, and average standard deviation (SD) for each lead, for each recording in the entire ECG dataset. We discarded any ECG containing a lead with an average QRS complex amplitude outside 2 standard deviations of the population QRS amplitude mean for that lead. We also discarded any ECG containing a lead with an SD outside 2 standard deviations of the population mean SD for that lead. **(Figure 1: CONSORT diagram)** The ECG waveforms that passed quality control for leads I, II, V1 – V6 were used for input into our model (see **Model Selection and Architecture)** and plotted as a two-dimensional image.

Patient age and ECG cart extracted parameters (Corrected QT interval, PR interval, Atrial Rate, and Ventricular Rate) were also acquired from XML files and utilized for input to our model. Overall distribution of input variables with respect to each outcome is shown in the pairplots of **Supplementary Figures 4 and 5**. We found that input variables were not correlated either with respect to each other, or the outcome.

Finally, no ECGs were excluded based upon associated diagnoses in the hope for generalizability across pathologies.

### Model selection and Architecture

ECG waveform data consisting of arrays of numbers (vectors) can be processed using either a 1D Convolutional Neural Network (CNN) or a 2D CNN. Typically, 2D CNNs are more computationally intensive, and are frequently used in image processing or genomics studies^46^. We elected to use a 2D CNN because all institutions may not store ECG data as vectors, and to be able to leverage pre-trained, robust 2D CNN architectures via transfer learning. We assessed different pre-built 2D CNN architectures at or about the same level of complexity as the backbone of our modeling (see **Supplementary Materials**) and identified the Efficientnet^47^ as the best performing CNN for our modelling task **(Supplementary Figure 6)**.

We created a unique multi-modal DL architecture used for our analyses. ECG extracted parameters and age are considered tabular data which form one part of the input, while the ECG data (image) form the other **(Supplementary Figure 7)**. Tabular data was processed through a Multi-Layer Perceptron (MLP) architecture, while imaging data was processed through the CNN architecture described above. The outputs of both the MLP and CNN were combined utilizing fully connected layers with ReLU activations, with each containing 128, 64, 32 and 16 neurons in the layers leading to the final layer.

For LVEF classification we utilized Cross-Entropy loss, with the final layer consisting of 3 neurons and a SoftMax activation function. For LVEF regression, we utilized L1 loss, with the final layer consisting of 1 neuron and a ReLU activation function. For RVD + RVSD, we utilized Binary Cross-Entropy loss, with the final layer consisting of one neuron, with a *Sigmoid* activation function. All models used the Adam optimizer with a learning rate of 1e-4.

### Experimental design

We collated data from four Mount Sinai facilities (Mount Sinai Hospital, Mount Sinai Brooklyn, Mount Sinai West, and Mount Sinai Beth Israel) to form an internal testing dataset. All data collected from Mount Sinai Morningside formed the external validation dataset. We confirmed that no patients within the external validation were present in the internal testing dataset. The relative distributions of the dataset across internal testing and external validation for each outcome are shown in **Tables 1 and 2**.

**Table 2:** Population metrics of the study population by Right Ventricular Systolic Dysfunction or Right Ventricular Dilation category

|  | RVSD or RVD | Normal | p |
| --- | --- | --- | --- |
| **Internal Testing Cohort** |  |  |  |
| Patients | 30,780 | 104436 |  |
| Echo-ECG pairs | 278877 | 425815 |  |
| Age | 65.8 (65.8 - 65.9) | 66.7 (66.7 - 66.8) | <0.0001 |
| Gender n (%) |  |  | <0.0001 |
| Male | 18,533 (60.21%) | 51122 (48.95) |  |
| Female | 12,247 (39.79%) | 53314 (51.05) |  |
| Race n (%) |  |  | <0.0001 |
| American Indian | 384 (1.25%) | 962 (0.92%) |  |
| Asian | 708 (2.3%) | 2414 (2.31%) |  |
| Black | 3,258 (10.58%) | 8621 (8.25%) |  |
| Hispanic | 2,478 (8.05%) | 6569 (6.29%) |  |
| Other | 2,513 (8.16%) | 12715 (12.17%) |  |
| Pacific Islander | 43 (0.14%) | 117 (0.11%) |  |
| Unknown | 12,300 (39.96%) | 44079 (42.21%) |  |
| White | 9,096 (29.55%) | 28959 (27.73%) |  |
| Ventricular Rate | 82.4 (82.2 - 82.6) | 77.8 (77.7 - 77.9) | <0.0001 |
| Atrial Rate | 99.1 (98.4 - 99.8) | 84.5 (84.2 - 84.8) | <0.0001 |
| PR Interval | 170.1 (169.7 - 170.5) | 163.8 (163.6 - 164.0) | <0.0001 |
| QTc Interval | 464.1 (463.6 - 464.7) | 446.2 (445.9 - 446.4) | <0.0001 |

| External Validation Cohort |  |  |  |
| --- | --- | --- | --- |
| Patients | 1,783 | 11,649 |  |
| Echo-ECG pairs | 8,828 | 47,990 |  |
| Age | 67.2 (66.9 - 67.5) | 67.3 (67.1 - 67.4) | 0.06 |
| Gender n (%) |  |  | <0.0001 |
| Male | 1,067 (59.84%) | 5,881 (50.49%) |  |
| Female | 716 (40.16%) | 5,768 (49.51%) |  |
| Race n (%) |  |  | <0.0001 |
| American Indian | - | - |  |
| Asian | - | 35 (0.3%) |  |
| Black | 289 (16.21%) | 1629 (13.98%) |  |
| Hispanic | 33 (1.85%) | 159 (1.36%) |  |
| Other | 502 (28.15%) | 3775 (32.41%) |  |
| Pacific Islander | - | - |  |
| Unknown | 587 (32.92%) | 3483 (29.9%) |  |
| White | 368 (20.64%) | 2564 (22.01%) |  |
| Ventricular Rate | 86.0 (85.0 - 86.9) | 79.9 (79.6 - 80.3) | <0.0001 |
| Atrial Rate | 104.5 (101.5 - 107.5) | 86.4 (85.6 - 87.2) | <0.0001 |
| PR Interval | 171.8 (170.0 - 173.6) | 166.9 (164.7 - 169.2) | <0.0001 |
| QTc Interval | 473.6 (471.5 - 475.8) | 451.9 (451.2 - 452.6) | <0.0001 |

### Model Training and Testing

We utilized a Group Stratified K-fold cross-validation design. This robust procedure involves treating each patient as a separate group and restricts them to either training or testing datasets, thereby preventing leakage of Echo-ECG pairs. Stratified K-fold cross validation entails training a model on K separate data splits each retaining the class distribution of the overall dataset. We used K=10 cross validation folds for each outcome in internal testing.

In order to account for minor temporal variations, each label corresponding to an echo report within training data was paired to all ECGs performed 7 days before or after the echo. For evaluation, we emulated real-world conditions by taking only one echo report per patient, and only considering the ECG closest to that echo within the ±7-day timeframe.

For each fold, 15% of the testing dataset was partitioned into an internal validation dataset. Following each round (i.e., epoch) of training, performance was evaluated on both the training dataset and this internal validation dataset. This procedure was done both to ensure that the model was learning the salient features of the input dataset, while also not overfitting to them. We used a custom early stopping module to interrupt the training loop when performance on the internal validation dataset stopped improving for five epochs. At this point, the model was evaluated on the remainder of the internal testing data, and the external validation data.

### Performance Evaluation

Model performance for classification tasks was primarily evaluated by utilizing Area Under Receiver Operating Characteristic *curve* (AUROC), and Area Under Precision Recall Curve (AUPRC) metrics. Additionally, we also considered the Youden J index^48^ for calculation of threshold dependent metrics. For the regression task, we utilized Mean Absolute Error (MAE) as the evaluation metric (See **Supplementary Materials** for a more detailed discussion of these metrics).

### Model Explainability

While Deep Neural Networks often suffer from lack of easy interpretability, it is possible to derive which features of the input pushed the model towards a prediction. We utilized the captum framework^49^ for explainability due to its support for multi-modal interpretability. Plots were created showing the region on the ECG plot that contributed most to the overall prediction, in addition to showing relative contributions of both the ECG plot, as well as the tabular data extracted from the ECG.

### Software and hardware

All analysis was performed using the pandas^50^, numpy^51^, scipy^52^, scikit-learn^53^, PIL^54^, torchvision^55^ and Pytorch^56^ libraries. Model explainability was derived utilizing captum^49^. Plotting was performed using the matplotlib^57^ and seaborn^58^ libraries. All program code was written for the Python programming language (3.9.x)^59^. All software was run within custom Docker^60^ containers created from official PyTorch docker images. Models were trained on a HIPAA-compliant Azure Cloud virtual machine containing 4x NVIDIA v100 GPUs with 16GB VRAM each.

## Results

### Performance of NLP algorithm for labeling RV abnormalities

We built a rule-based NLP algorithm in order to identify RVSD and RVD outcomes from echo reports. To assess validity of this procedure, human generated labels for these reports were compared to algorithm generated labels and quantified in terms of correctly classified labels, incorrectly classified labels, as well as missed labels.

From 420 outcomes included in review for RV function, we correctly classified 404, did not predict a label for 13, and incorrectly classified 3. For RV size, from the 420 outcomes included in review, we correctly classified 402 outcomes, did not predict a label for 17, and incorrectly classified 1. Within detected outcomes, we achieved an overall accuracy of 99.7% for extraction of either RV Size or RV Function **(Supplementary Table 2)**.

### Performance of LVEF classification

We built a machine learning model to classify LVEF in terms of the following clinically relevant categories: <=40%, >40% and <=50%, as well as >50% from an ECG **(Table 3)**. We provide the outcome distribution for the LVEF dataset and experiments in **Table 1** and in the pairplot in **Supplementary Figure 4**.

**Table 3:**
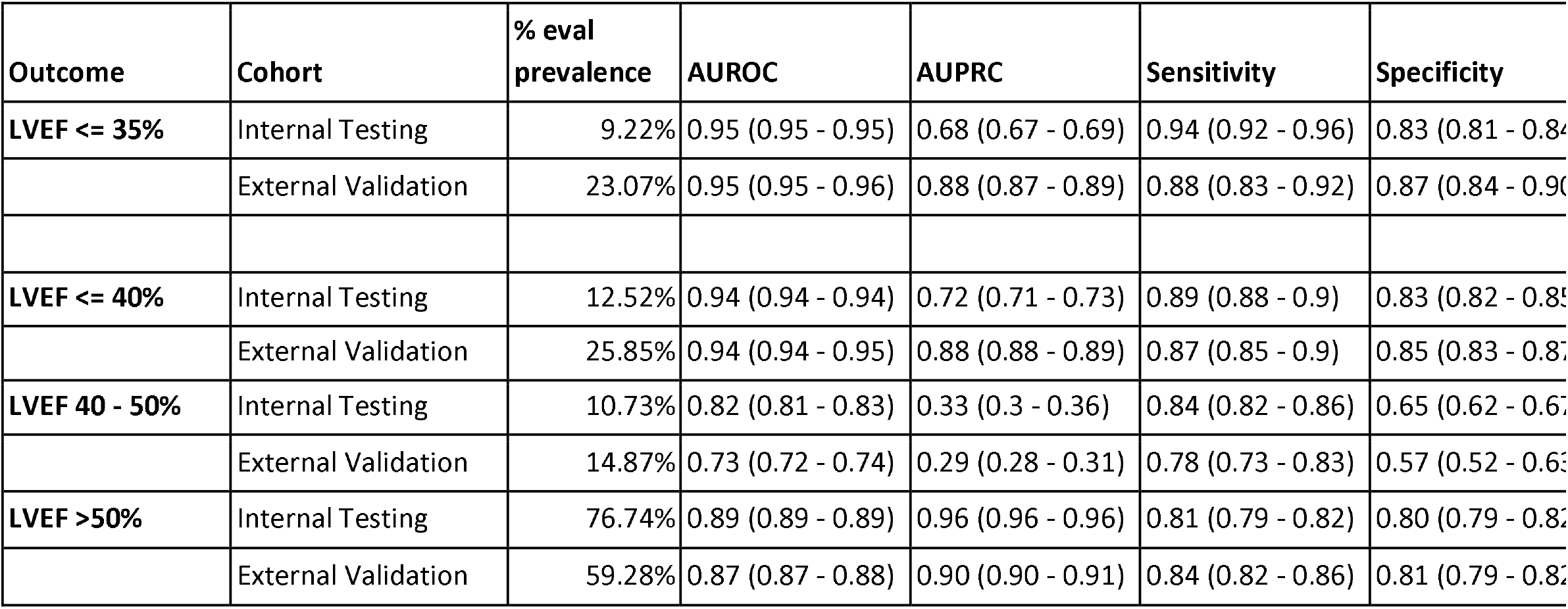
Performance at LVEF classification. Sensitivity and Specificity have been derived using the Youden J index.

| Outcome | Cohort | % eval prevalence | AUROC | AUPRC | Sensitivity | Specificity |
| --- | --- | --- | --- | --- | --- | --- |
| **LVEF <= 35%** | Internal Testing | 9.22% | 0.95 (0.95 - 0.95) | 0.68 (0.67 - 0.69) | 0.94 (0.92 - 0.96) | 0.83 (0.81 - 0.84) |
|  | External Validation | 23.07% | 0.95 (0.95 - 0.96) | 0.88 (0.87 - 0.89) | 0.88 (0.83 - 0.92) | 0.87 (0.84 - 0.90) |
| **LVEF <= 40%** | Internal Testing | 12.52% | 0.94 (0.94 - 0.94) | 0.72 (0.71 - 0.73) | 0.89 (0.88 - 0.9) | 0.83 (0.82 - 0.85) |
|  | External Validation | 25.85% | 0.94 (0.94 - 0.95) | 0.88 (0.88 - 0.89) | 0.87 (0.85 - 0.9) | 0.85 (0.83 - 0.87) |
| **LVEF 40 - 50%** | Internal Testing | 10.73% | 0.82 (0.81 - 0.83) | 0.33 (0.3 - 0.36) | 0.84 (0.82 - 0.86) | 0.65 (0.62 - 0.67) |
|  | External Validation | 14.87% | 0.73 (0.72 - 0.74) | 0.29 (0.28 - 0.31) | 0.78 (0.73 - 0.83) | 0.57 (0.52 - 0.63) |
| **LVEF >50%** | Internal Testing | 76.74% | 0.89 (0.89 - 0.89) | 0.96 (0.96 - 0.96) | 0.81 (0.79 - 0.82) | 0.80 (0.79 - 0.82) |
|  | External Validation | 59.28% | 0.87 (0.87 - 0.88) | 0.90 (0.90 - 0.91) | 0.84 (0.82 - 0.86) | 0.81 (0.79 - 0.82) |

Our model performed extremely well at detecting patients with an LVEF of <= 40% both for internal testing (12.52% prevalence) and external validation (25.85% prevalence) with AUROC values of 0.94 (95% CI: 0.94 - 0.95) in each case. This trend was maintained for the Precision Recall Curves as well, with AUPRC values of 0.72 (95% CI: 0.71 - 0.73) for internal testing, going up to 0.88 (95% CI: 0.88 - 0.89) in external validation.

Similar results were seen for detection of LVEF > 50%. For internal testing (76.7% prevalence), the model achieved an AUROC of 0.89 (95% CI: 0.89 - 0.89), which was maintained for external validation (59.3% prevalence) at 0.87 (95% CI: 0.87 - 0.88). AUPRC values were also exceptional at 0.96 (95% CI: 0.96 - 0.96) for internal testing and 0.90 (95% CI: 0.90 - 0.91) for external validation.

Performance was lower for LVEF values >40% and <=50%. In the internal testing dataset (10.73% prevalence), we achieved an AUROC of 0.82 (95% CI: 0.81 - 0.83), and for external validation, this value was 0.73 (95% CI: 0.72 - 0.74). AUPRC values were 0.33 (95% CI: 0.3 - 0.36) for the testing dataset, and 0.29 (95% CI: 0.28 - 0.31) for the external validation dataset.

We show ROC curves in **Figure 2**, and PR curves in **Supplementary Figure 8**. The interpretability plots for LVEF prediction **(Figure 3)** using our explainability framework highlighted QRS complexes for prediction of each LVEF related outcome. The relative importance of the extracted features was found to be variable across tested patients.

**Figure 2:**
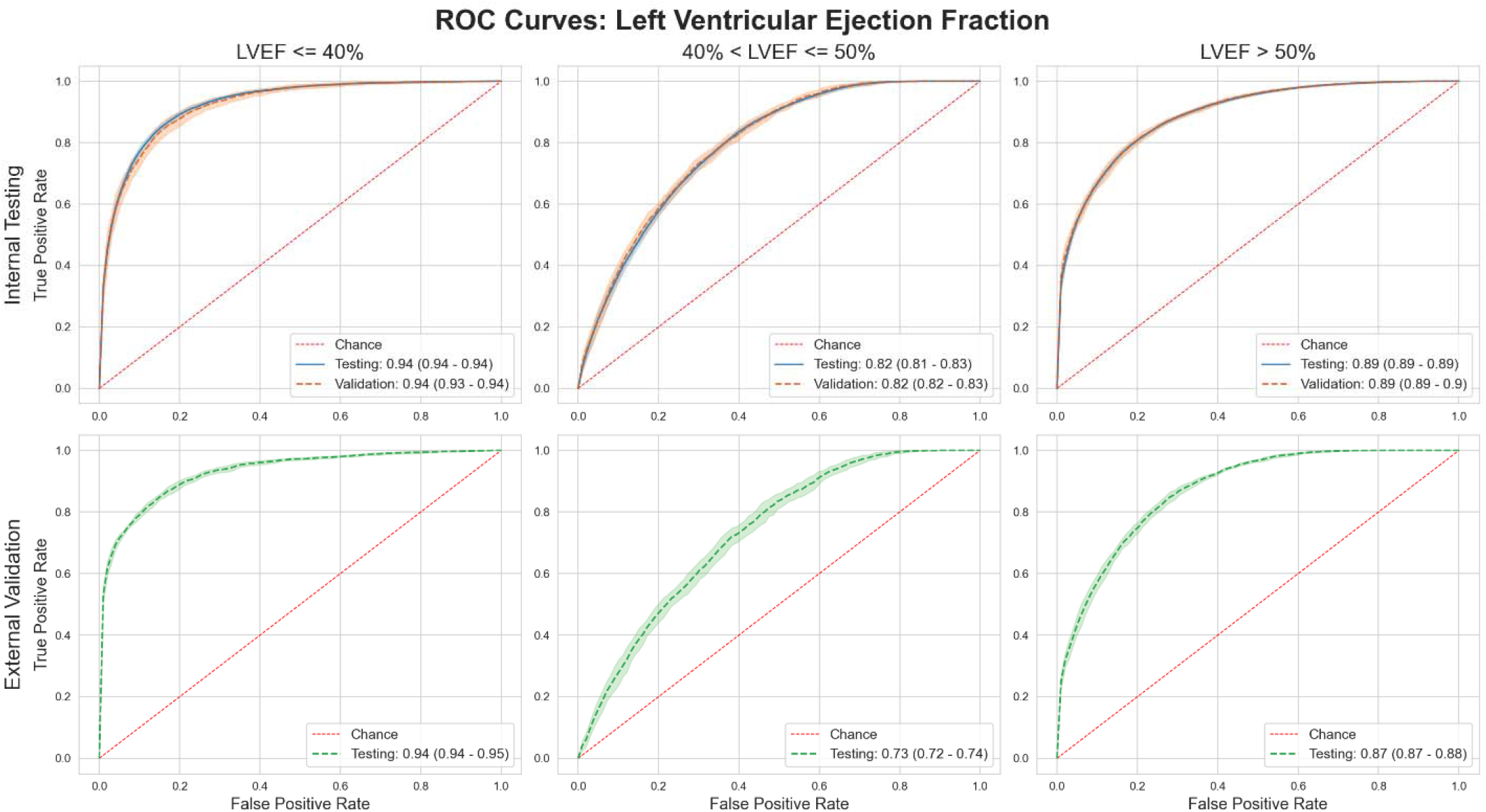
Receiver Operating Characteristic Curves: LVEF Classification. Upper row shows performance for each outcome in the Internal Testing cohort, while the lower row shows performance in the external validation cohort.

**Figure 3:**
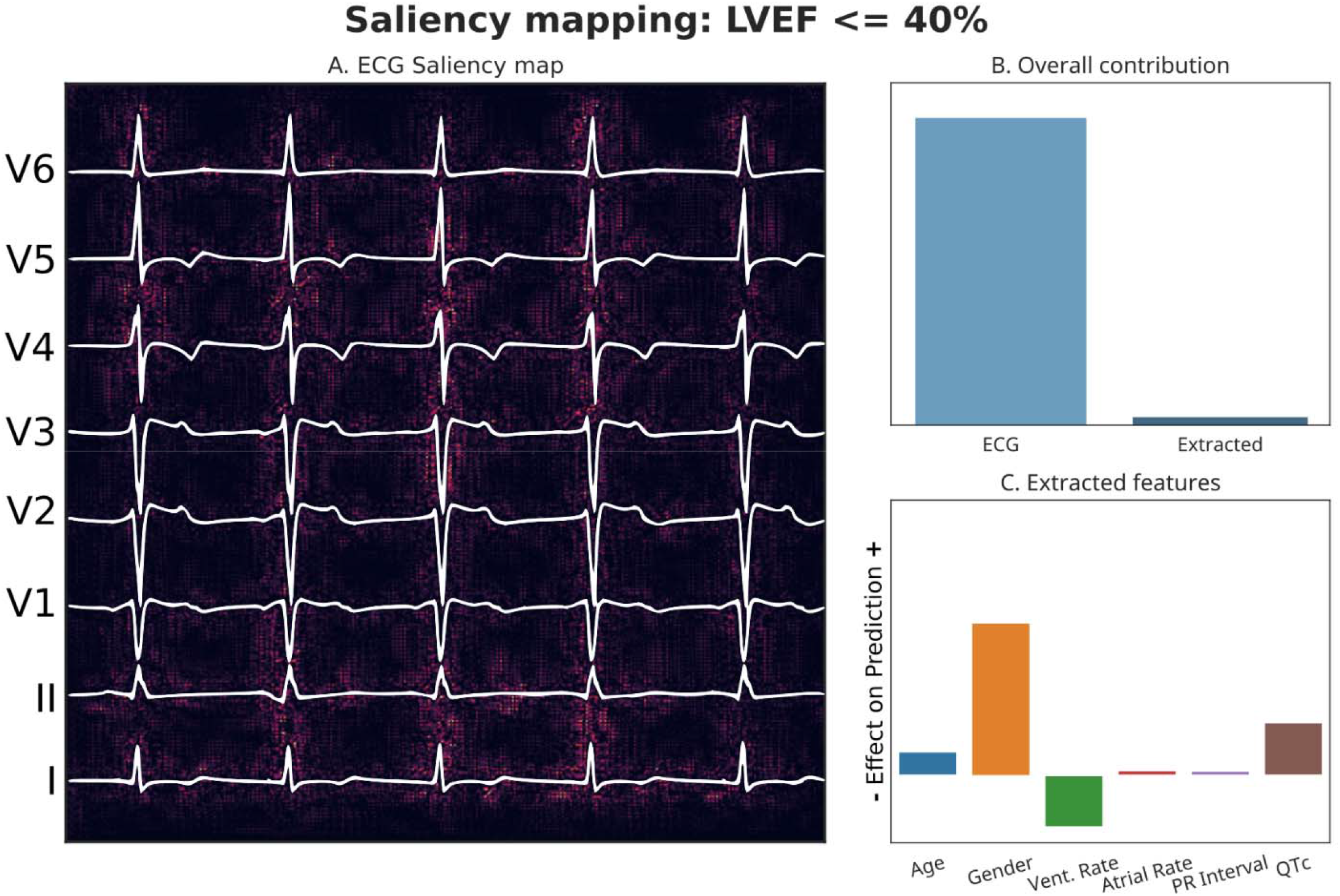
LVEF Explainability. Panel A: Pixels of input image which were most responsible for driving the prediction towards an LVEF of <40% are highlighted. Panel B: Relative contributions of imaging data and tabular data to the overall prediction. Panel C: Effect of the tabular features on model’s prediction. *Predicted LVEF <40% probability: 0.89. Actual LVEF value: 29%*

Model performance was maintained when tested against varying severity of MR, with better performance seen when tested against Normal - Mild MR. **(Supplementary Table 3, Supplementary Figures 9, 10)**

In a separate experiment for detection of patients with an LVEF <= 35%, our model performed exceedingly well in internal testing (9.22% prevalence), with an AUROC of 0.95 (95% CI: 0.95 – 0.95), and an AUPRC of 0.68 (95% CI: 0.67 – 0.69). In external validation (23.07% prevalence), these results were maintained with an AUROC of 0.95 (95% CI: 0.95 – 0.95 AUPRC of 0.88 (95% CI: 0.87 – 0.89). **(Table 3, Supplementary Figures 11, 12)**

### Performance of LVEF Regression

We constructed a deep learning model to predict the exact value of LVEF from an Echo-ECG pair within a regression framework. Within the internal testing dataset, the MAE was 5.84% (95% CI: 5.82 - 5.85%). For the external validation dataset, the MAE was 6.14% (95% CI: 6.13 - 6.16%). A scatterplot showing the relationship between predicted and actual values for the overall dataset is shown in **Figure 4**.

**Figure 4:**
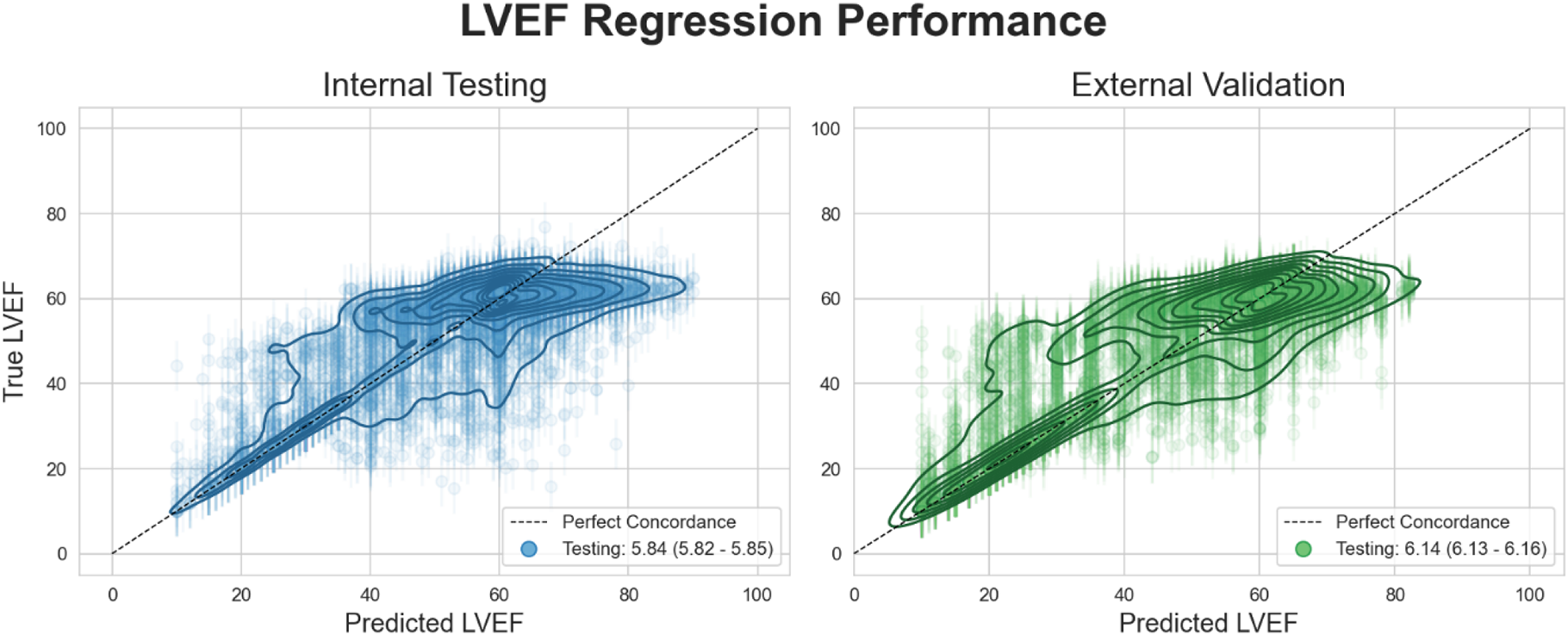
Scatterplot showing LVEF regression performance. Contour lines show density of predicted LVEF values vs ground truth LVEF values around the line of perfect concordance. Error bars around each datapoint indicate the mean absolute error. 10,000 samples shown for each panel to prevent overplotting. Contour maps were generated using the entire dataset.

We evaluated the performance of this regression model within clinically relevant LVEF subgroups. In the first subgroup of echo derived LVEF <=40%, the MAE for the model was 6.69% in internal testing, and 6.46% in external validation. Within the second subgroup of LVEF between 40 and 50%, the MAE was greater at 8.08% in internal testing, and 8.55% in external validation. Within the final subgroup of patients of LVEF > 50%, our model achieved a MAE of 5.41% in internal testing, and 5.44% in external validation **(Supplementary Figure 13)**.

### Performance of Right Ventricular Systolic Dysfunction and Right Ventricular Dilation classification

We built a deep learning model to predict either RVSD or RVD from a patient’s ECG in both internal testing (32.44% prevalence) as well as external validation (15.53% prevalence) **(Table 2)**. Our model achieved robust performance in this task with an AUROC of 0.84 (95% CI: 0.84 - 0.84) in internal testing, maintained in external validation at 0.84 (95% CI: 0.84 - 0.84) **(Figure 5, Table 4)**. Our model achieved similar success with respect to AUPRC, with values of 0.67 (95% CI: 0.66 - 0.67) in testing, and 0.55 (95% CI: 0.54 - 0.55) in external validation. **(Supplementary Figure 14, Table 4)**. Plots created using the explainability framework again highlighted QRS complexes for prediction of the composite outcome. **(Figure 6)**.

**Table 4:**
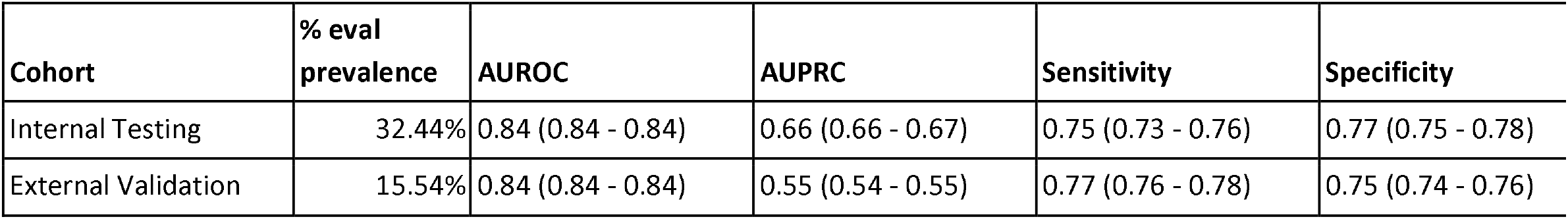
Performance at RVSD + RVD composite outcome classification. Sensitivity and Specificity have been derived using the Youden J index.

**Figure 5:**
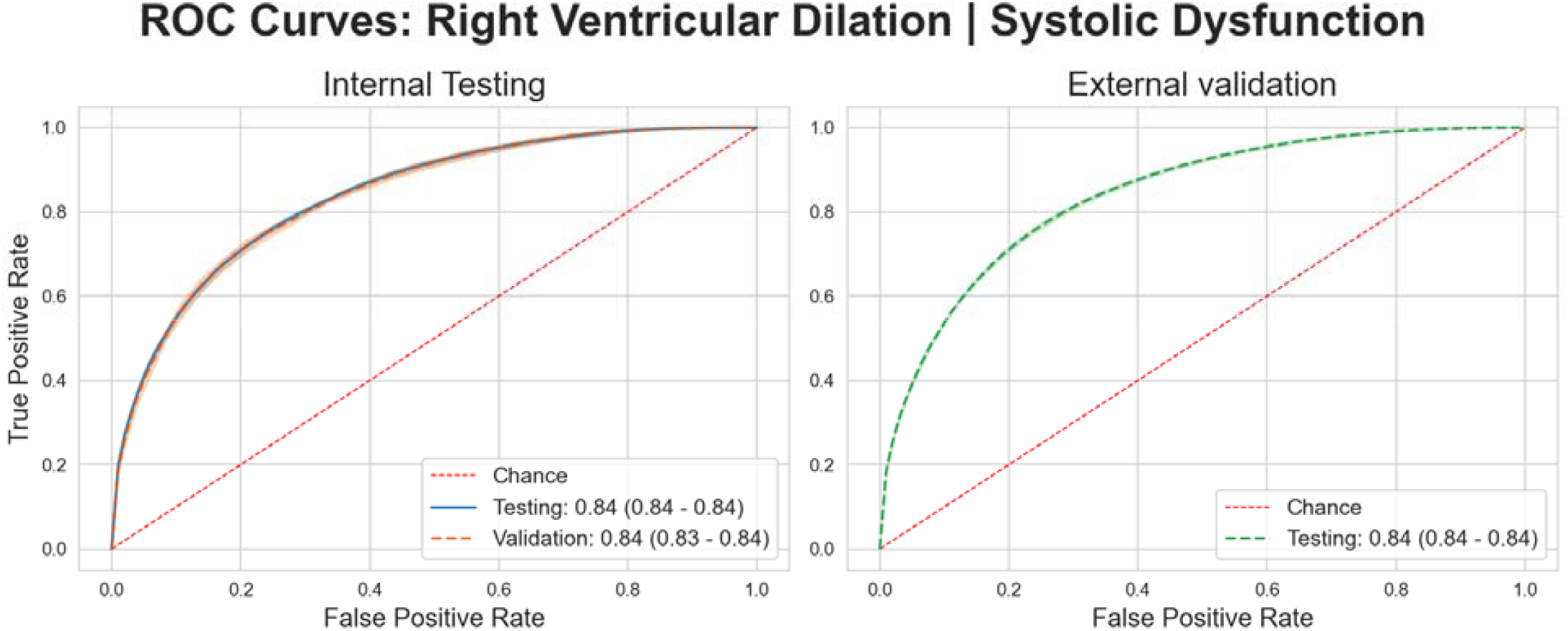
Receiver Operating Characteristic Curves: Right Ventricular Dilation or Right Ventricular Systolic Dysfunction classification. Left panel shows performance for the Internal Testing cohort, while the right panel shows performance in the external validation cohort.

**Figure 6:**
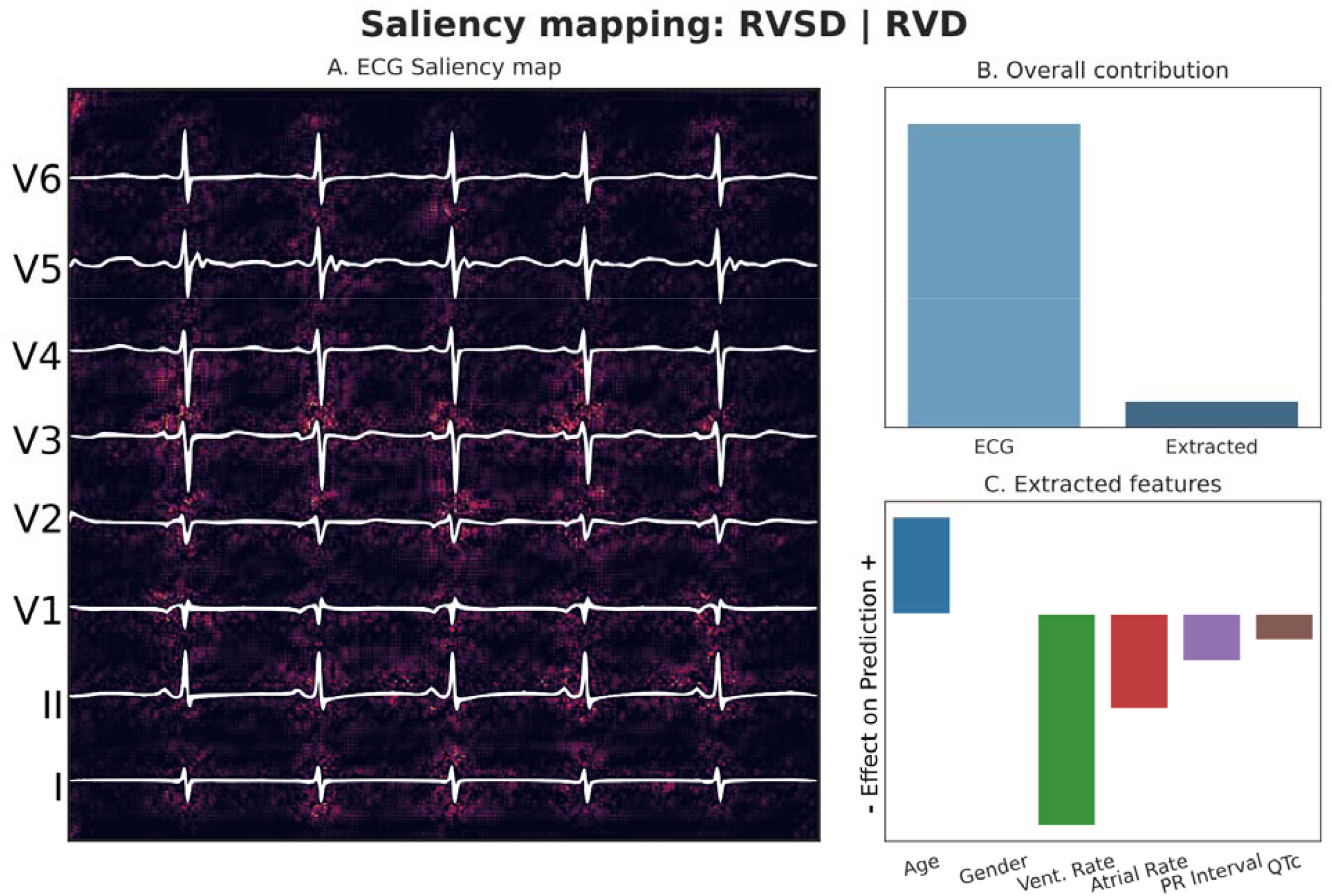
RV Composite Outcome (RVSD or RVD) Explainability. Panel A: Pixels of input image which were most responsible for driving the prediction towards the composite outcome are highlighted. Panel B: Relative contributions of imaging data and tabular data to the overall prediction. Panel C: Effect of the tabular features on model’s prediction. *Predicted probability of composite outcome: 0.948, RVSD: Present, RVD: Present*

### Performance of Right Ventricular Systolic Dysfunction and Right Ventricular Dilation Prediction with Variations in LVEF

We next performed a subgroup analysis to assess model performance in the presence of very low (<=40%) or close to normal (>50%) LVEF. In the internal testing cohort (<=40% prevalence: 70.93%, >50% prevalence: 19.49%), AUROC was similar, albeit slightly lower at values of 0.81 (95% CI: 0.79 - 0.82) for LVEF <=40%, and 0.82 (0.81 - 0.83) for LVEF > 50%. This performance decrease was not seen in external validation, with AUROC values of 0.89 (95% CI: 0.88 - 0.89) for LVEF <=40%, and 0.85 (95% CI: 0.84 – 0.86) for LVEF >50%. **(Supplementary Figure 15, Supplementary Table 4)**

Interestingly, there was a much more profound variation in the AUPRC values derived from this analysis. The AUPRC was enhanced at 0.91 (95% CI: 0.9 - 0.92) in the presence of an LVEF <= 40%, while persisting at 0.57 for LVEF > 50%. This pattern was also seen in external validation where the AUPRC was 0.92 (95% CI: 0.91 - 0.93) for LVEF <=40%, and 0.63 (95% CI: 0.59 - 0.67) for LVEF >50%. **(Supplementary Figure 16, Supplementary Table 4**)

## Discussion

Utilizing 700,000 ECGs for around 150,000 unique patients from a large and socioeconomically diverse cohort of patients in the Mount Sinai Health System in New York City. We developed, evaluated, and externally validated multimodal deep learning models capable of discerning the contractile state of both the left and right ventricles. We created an accurate Natural Language Processing pipeline for extraction of outcomes from free text echo reports. Finally, we developed a multimodal explainability framework to highlight which parts of the ECG are more salient for each outcome and derive interactions between demographic data and imaging data.

Existing work on LVEF extraction from ECGs is limited to classification of LVEF values of <= 35%. We extend our classification framework to clinically pertinent ranges^42^ of <= 40%, 40 - 50%, and >50% to be able to catch additional downstream management and prognostic implications. For example, the difference between an LVEF of 41% and one of 71% is hemodynamically and clinically significant. We also outperform existing work at detection of LVEF <= 35%^40^ (AUROC: 0.93 vs 0.95), in a much more ethnically and socioeconomically diverse set of patients. To the best of our knowledge, there has also been no work till date on estimating LVEF value as a continuous (percentage) number using ECGs. Clinical guidelines which segment patients based on LVEF assume that one set of classification boundaries is applicable to the entire population. However, normal variation in echo derived baseline values is expected secondary to patient demographics. A regression approach reduces risk of such misclassification^37^. We posit that LVEF regression may dramatically enhance the value of a screening ECG even in otherwise low risk groups and is also more useful for evaluation of LVEF in the longitudinal setting, where LVEF changes over time are as clinically important^61^. Additionally, the regression framework is also independent of changes in management guidelines.

Internal validation alone cannot guarantee model quality^58^. Biases within training data which help performance may not translate to external cohorts. It follows that external validation is extremely important to assess how generalizable a model is. We were encouraged to see that for evaluation of LVEF, there was minimal to low change in performance in going from internal to external validation.

Diagnosis of RV dysfunction using deep learning on ECGs is a novel approach. Our models perform robustly for the detection of the composite RVSD and RVD outcome. Additionally, the exceptionally high AUPRC values in the presence of LVEF <= 40% indicate such models are suited for tracking RV involvement secondary to HFrEF. Once again, the models translated extremely well to external validation. Our decision to not stratify RV disease according to severity was made to allow for early disease detection. Depending on clinical context, this may be adjusted to more severe disease. We posit performance in this context will also increase due to there being a greater difference between normal and diseased cases.

Deep learning represents a powerful set of tools that can find patterns that are too subtle for human perception. It has been applied with great success in natural image classification, and we have extended those capabilities into successfully reading ECGs for outcomes that do not have an established set of guidelines.

## Limitations

LVEF derived from an echo may vary between operators^62^, between tests, with use of echogenic contrast media, and even with range of reported LVEF. Echo operators find it easier to tell when an LVEF is very abnormal (<40%), or closer to normal (>50%), but more poorly in between those ranges. The higher reporting error in this range may lower performance, even if the model makes correct predictions. Overall, these issues are amenable to methods which lead to more accurate ground truth about the LVEF, such as cardiac MRIs, or universal use of echogenic contrast medium. Additionally, we paired ECGs to echo reports over a ±7-day time period. Changes in LVEF secondary to either acute pathology or treatment may lead to discordance between ground truth and recorded values. This may be offset by the resilience of neural network architectures to random error, as is evidenced by the robust performance of our models in either evaluation cohort. Additional random error may have been introduced by the NLP pipeline creating erroneous labels in some cases, although our accuracy upon manual review was very high (99.7%). External validity, though demonstrated at another hospital site, is still limited as the external site was still a part of the Mount Sinai health system and similar geographical location, further creating the need for future work to focus on prospective and greater site validation of our models.

## Conclusions

Deep learning models can extract information about biventricular function from the ECG that would ordinarily require an echocardiogram. Such models may enhance the usefulness of the ECG in the screening and management of either sided heart failure progressing to biventricular disease.

## Clinical Perspectives

### Competency in Systems-based Practice

Deep learning on electrocardiograms can simultaneously detect right ventricular systolic dysfunction or dilation and quantitatively estimate left ventricular ejection fraction.

### Translational Outlook

A prospective trial is needed to determine how automated deep learning tools on the electrocardiogram may be implemented to support clinical decision making in management of either sided heart failure progressing to biventricular disease

## Data Availability

Data utilized for training models cannot be released due to privacy concerns.

## Acknowledgements

The authors would like to thank Manbir Singh, Mark Shervey, Adeyinka Fadehan, Sparshdeep Kaur, Ejaz Siddiqui, Percy LaRosa, and Thareesh Dondapati for their invaluable support.

