## Supplementary Materials for "Using deep learning algorithms to simultaneously identify right and left ventricular dysfunction from the electrocardiogram"

Supplementary Methods

##### **Model architecture selection**

While the Efficientnet has been proven to perform better on natural image classification, we assessed whether it would similarly perform equally well on ECG data. Accuracy in deep learning is dependent on several factors. These include input scaling, and model complexity. Such concerns must also be balanced against computational budgets and wall time.

We sampled a random subset of 20,000 Echo-ECG pairs spread across all Mount Sinai facilities for training and evaluation with 3-fold group stratified cross validation. AUROC and AUPRC metrics were utilized to compare performance at quantification of LVEF <= 35%.

##### **Performance Metrics**

The Receiver Operating Characteristic Curve describes the relationship between the True Positive Rate (Sensitivity), and the False Positive Rate (1 - Specificity) at different probability thresholds. The area under this curve, or AUROC, ranges from 0 - 1, with higher values indicative of better performance. Specifically, this metric indicates the model’s discriminative ability, or its ability to tell positives from negatives.

The Precision Recall curve indicates the interplay between Precision (Positive Predictive Value) and Recall (Sensitivity) at different probability thresholds. The area under this curve, or AUPRC measures the quality of positive predictions, and is a better metric of model performance in more imbalanced datasets. AUPRC also ranges from the 0 - 1, with higher values being better.

The MAE is the sum of the absolute difference between true and predicted values, averaged over the number of observations in the dataset. The MAE ranges from 0 - positive infinity, with lower values representing less difference between the predicted and actual values. In other words, how divergent the model’s predictions were from the ground truth on average.

##### **Classification Thresholding using the Youden Index**

We report threshold dependent metrics derived from the Youden index. This selection of thresholds optimizes for both sensitivity and specificity. Since having high sensitivity is considered ideal for a screening tool, the threshold may be varied according to the potential cost of missing a patient vs the cost of a more rigorous follow up of a false positive.

Supplementary Results

##### **Model Architecture Selection**

The Efficientnet was found to have better performance at classification of LVEF <= 35%, while retaining a slight speed advantage of a few minutes per epoch. Following from this, we identified the Efficientnet architecture as the best performing model to use for this study. **(Supplementary Figure 6).** Efficientnets are a recent class of models known to attain greater performance (both accuracy and speed) through simultaneous balanced increases in complexity and input image size.

Supplementary Figures

**Supplementary Figures 1, 2, 3**: Annotated Echo reports

Phrases related to RV size, RV function, and Mitral valve status are each colored distinctly. Solid borders denote a diagnosis of “normal”, while dashed borders signify the presence of a pathology. Bold text within each annotation indicates the NLP derived diagnosis.

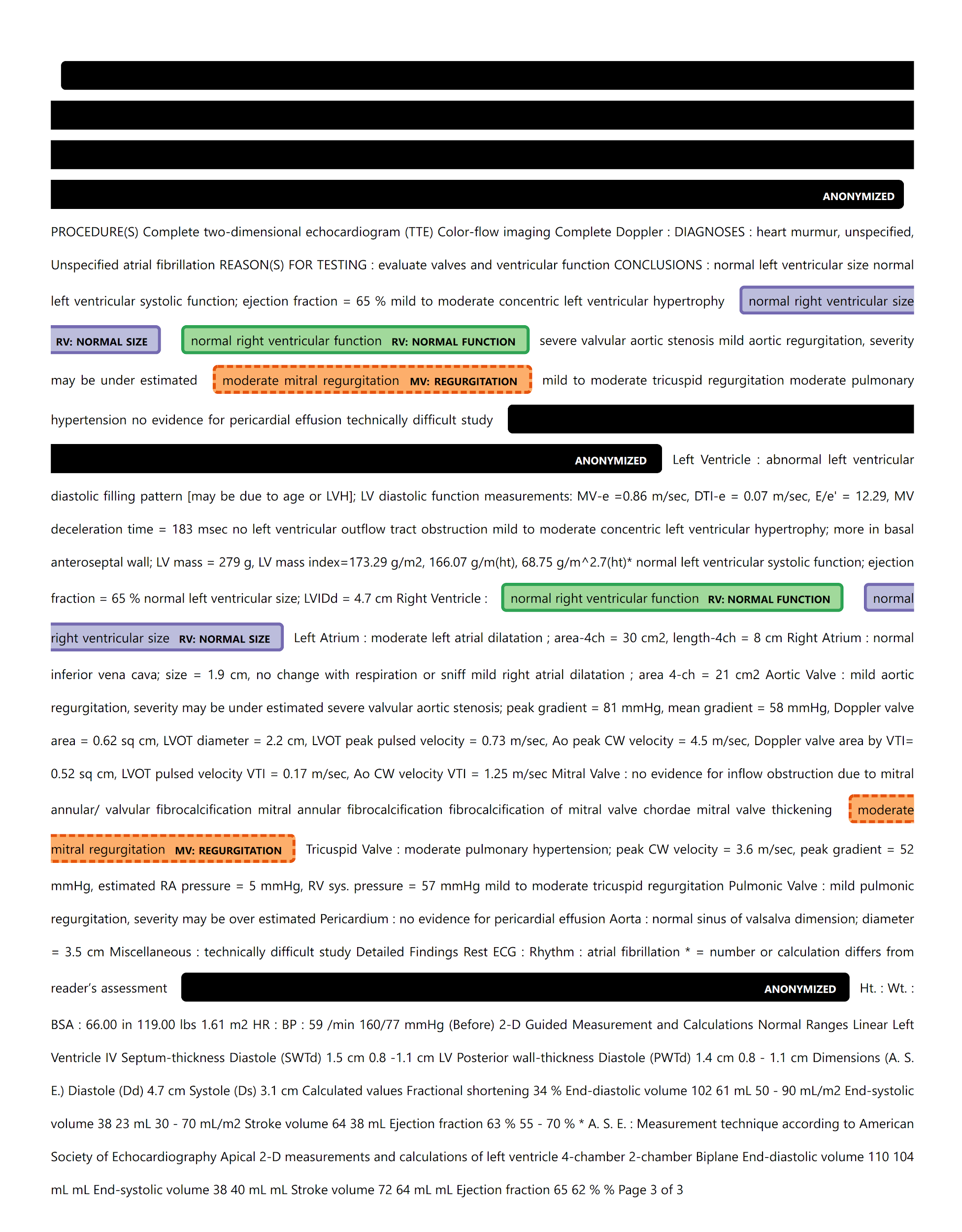

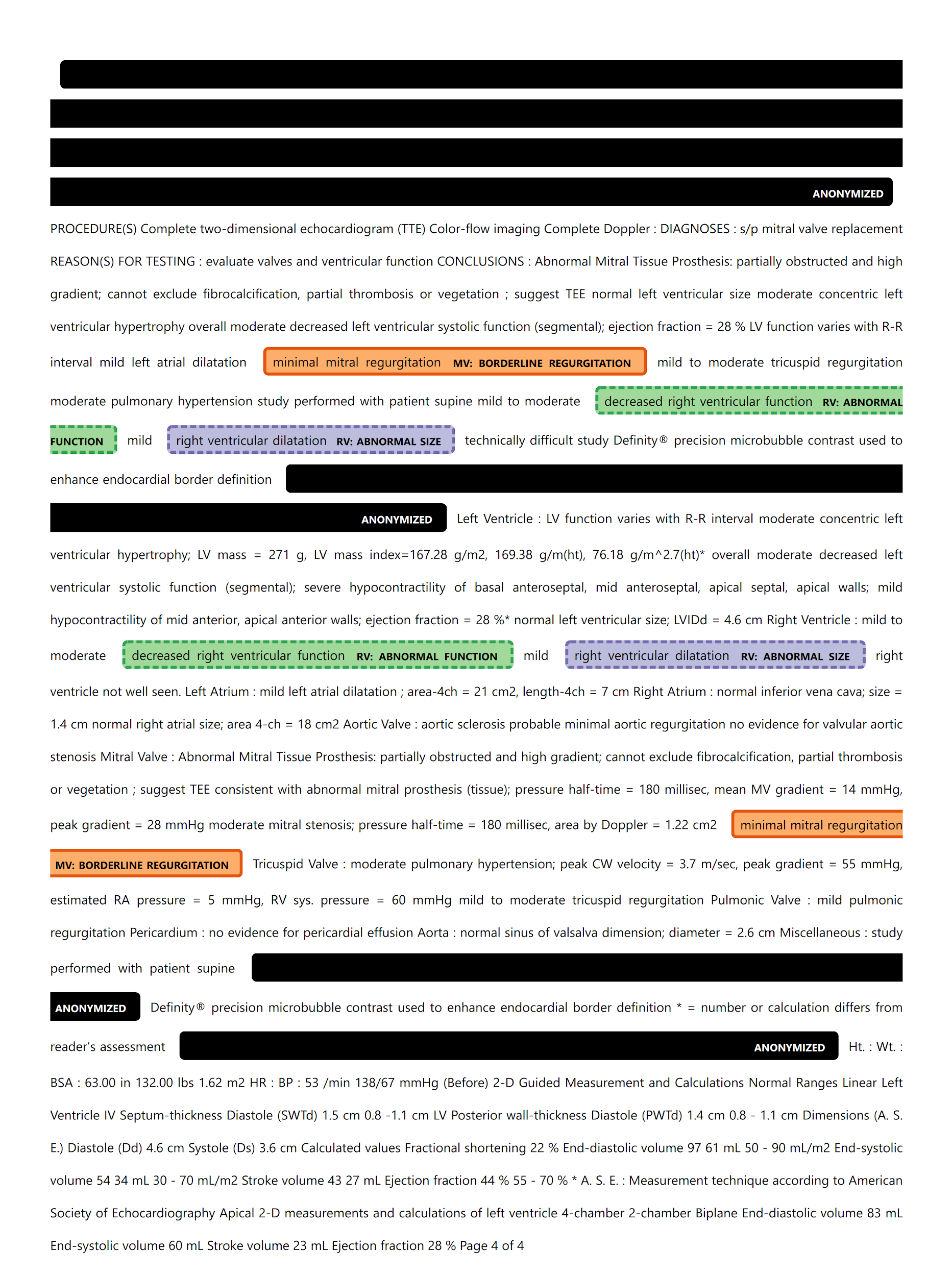

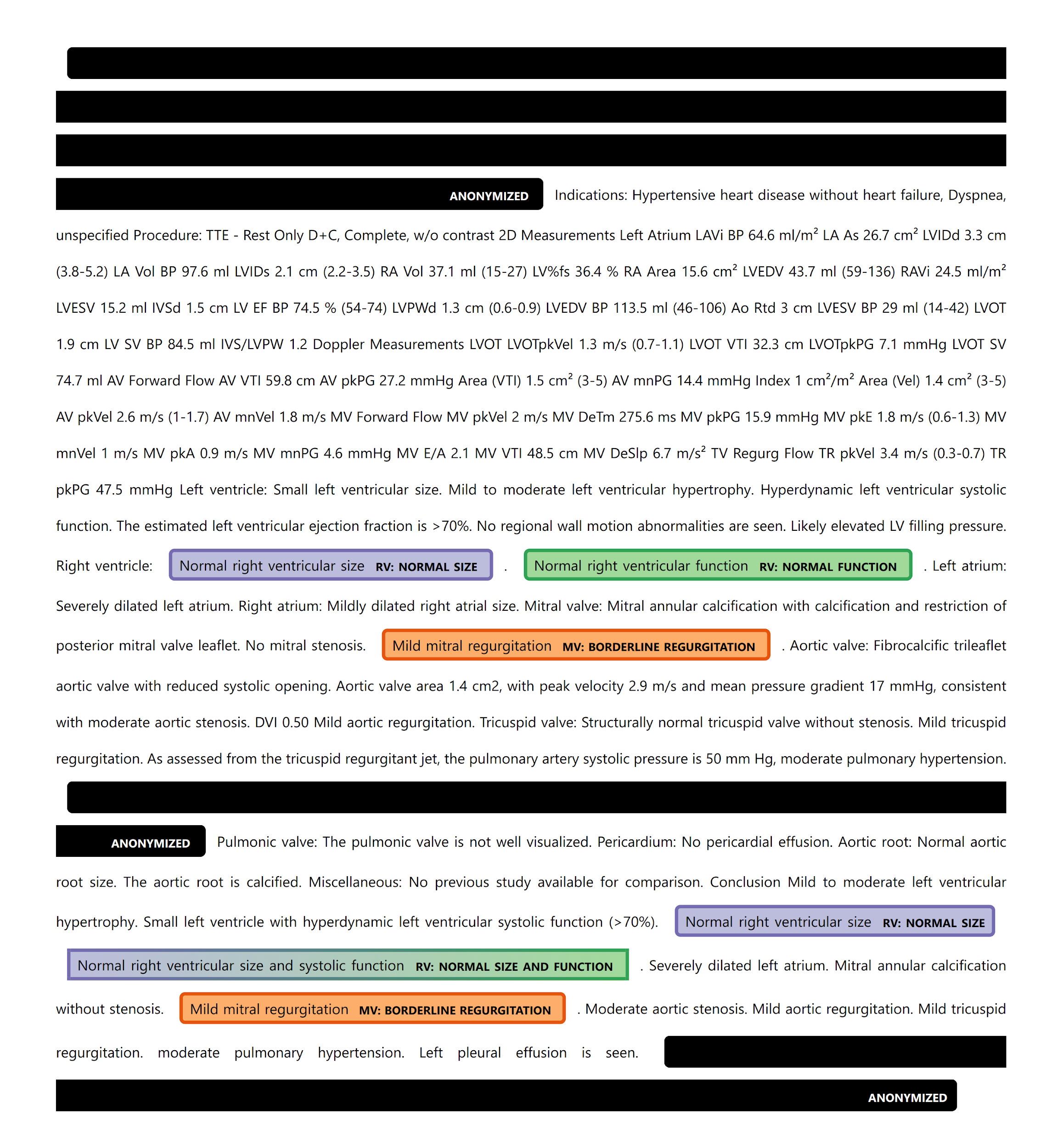

**Supplementary Figure 4: Left ventricular Ejection Fraction pairplot**

Pairplot showing relationship between values ECG extracted parameters, time delta between ECG and Echo, age, and ranges of LVEF. Each datapoint falling within a range is plotted in its own color. Random sampling (n=2000) was performed on the dataset while maintaining population distribution to prevent overplotting. Plots outside the diagonal are scatter plots showing the relationship between any two variables. Plots above the diagonal show topographic distribution of data points. The diagonal shows the Kernel Density Estimate of each LVEF category and overlaps between ranges are artifacts of extrapolation.

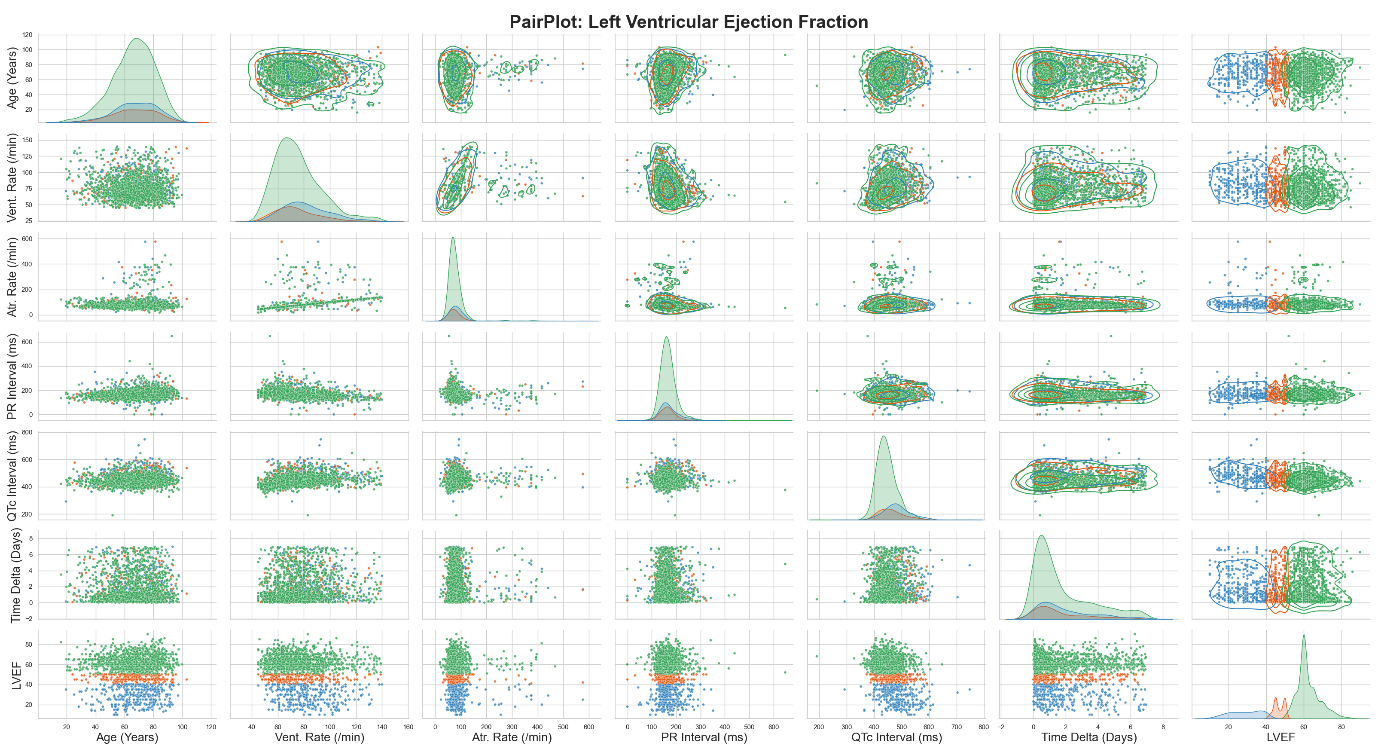

**Supplementary Figure 5: RVSD or RVD pairplot**

Pairplot showing relationship between values ECG extracted parameters, time delta between ECG and Echo, age, and composite RVSD + RVD outcome. Each data point corresponding to an outcome is shown in its own color. Random sampling (n=2000) was performed on the dataset while maintaining population distribution to prevent overplotting. Plots outside the diagonal are scatter plots showing the relationship between any two variables. Plots above the diagonal show topographic distribution of data points.

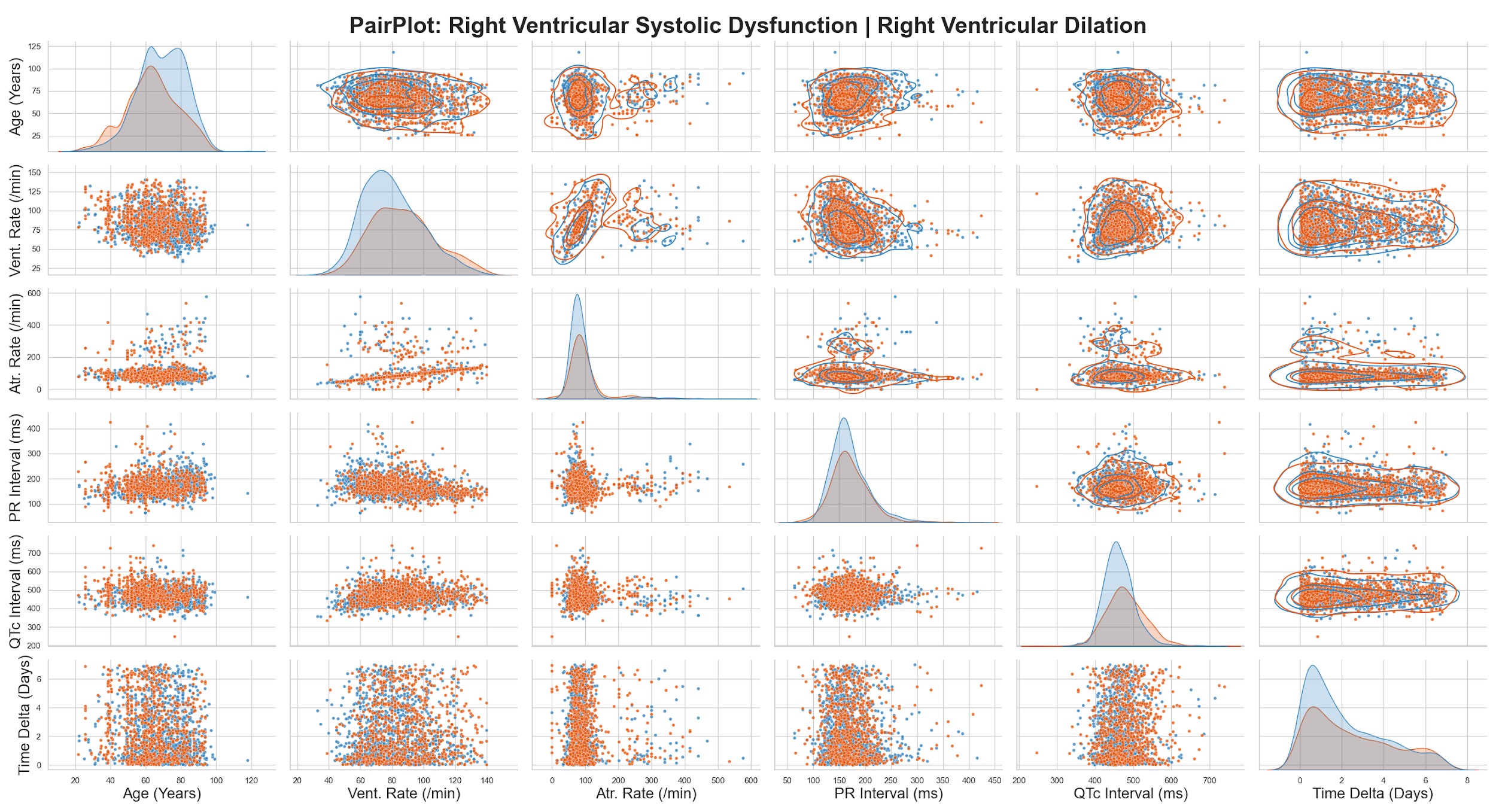

### **Supplementary Figure 6:** Performance comparisons between models of equivalent computational cost and complexity. Compared models include a Densenet201, an Efficientnet B4 architecture, and a Resnet 50.

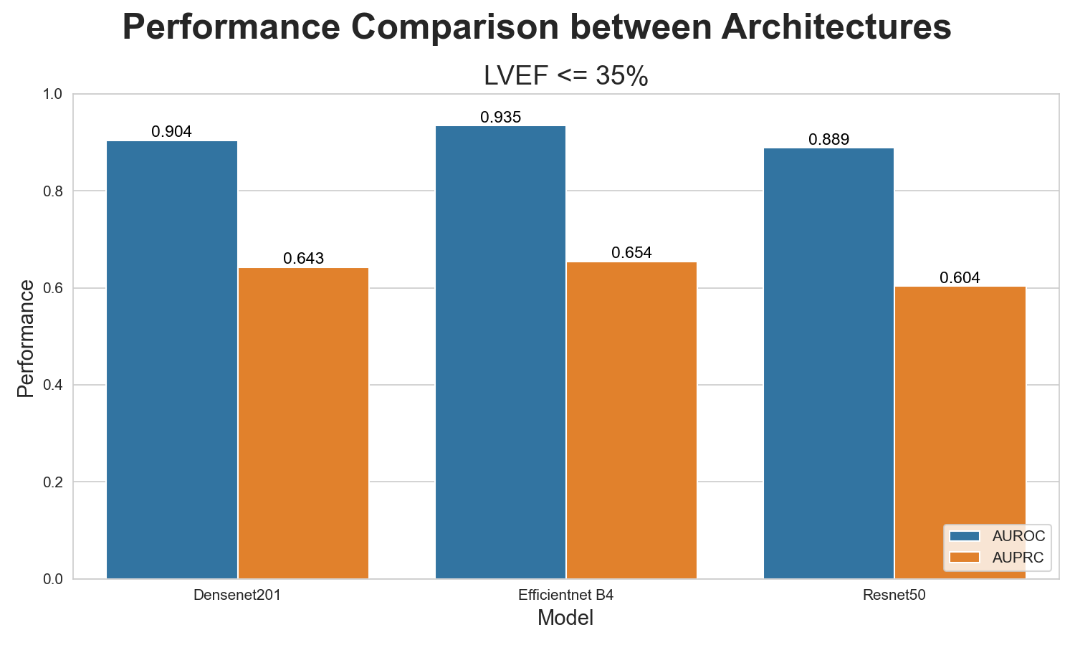

**Supplementary Figure 7**: Neural Network Architecture
Tabular (Extracted) data is input into the fully connected layers in Branch A, while imaging (ECG) data is input into the CNN in Branch B. Intermediate layers in (C) create a composite input from both the tabular data and CNN for the final output layer (D). Activations and shape of the output layer depend on the outcome. *ReLU: Rectified Linear Unit. SM: Softmax.*

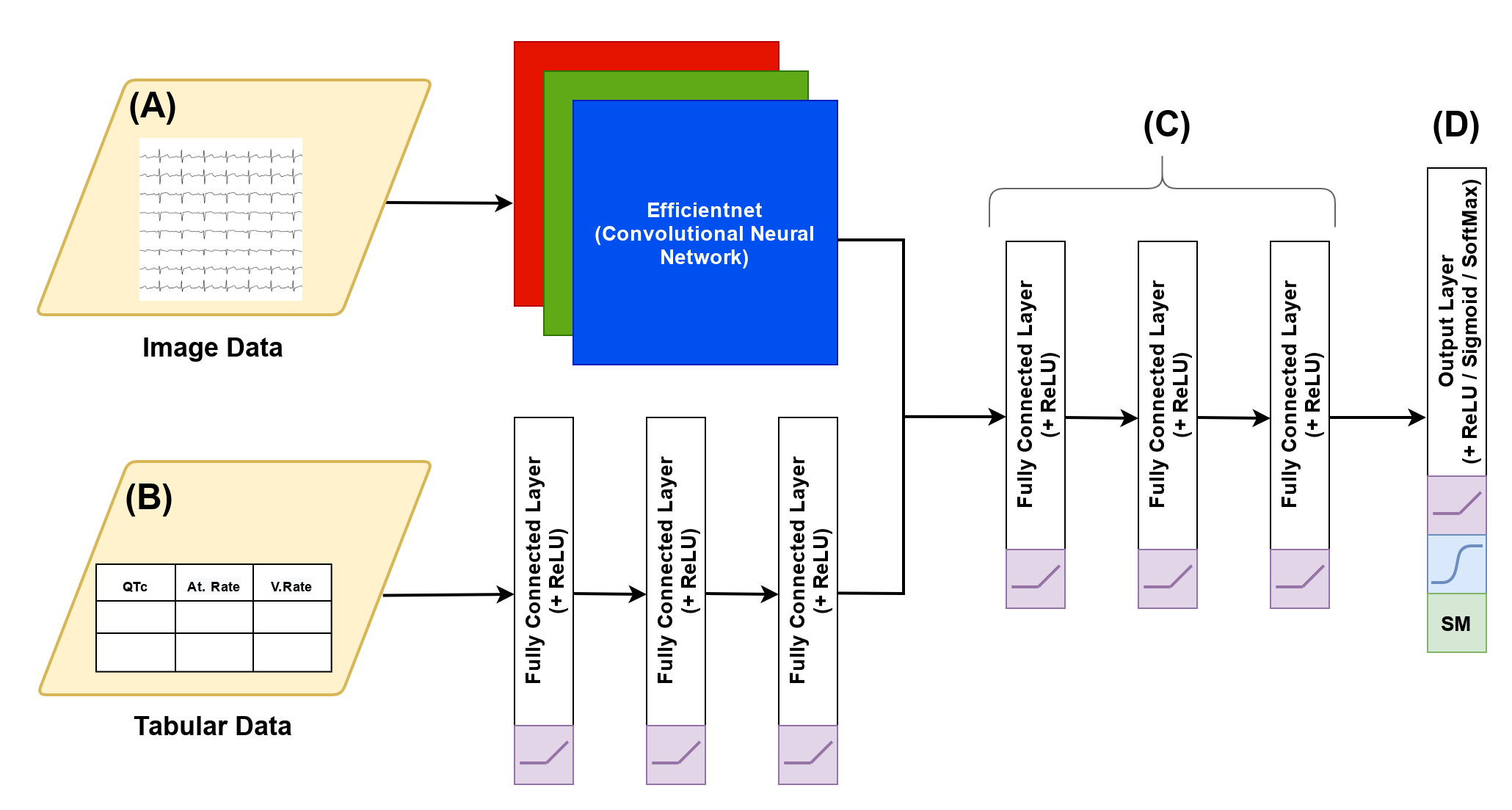

**Supplementary Figure 8**: Precision Recall Curves for LVEF classification

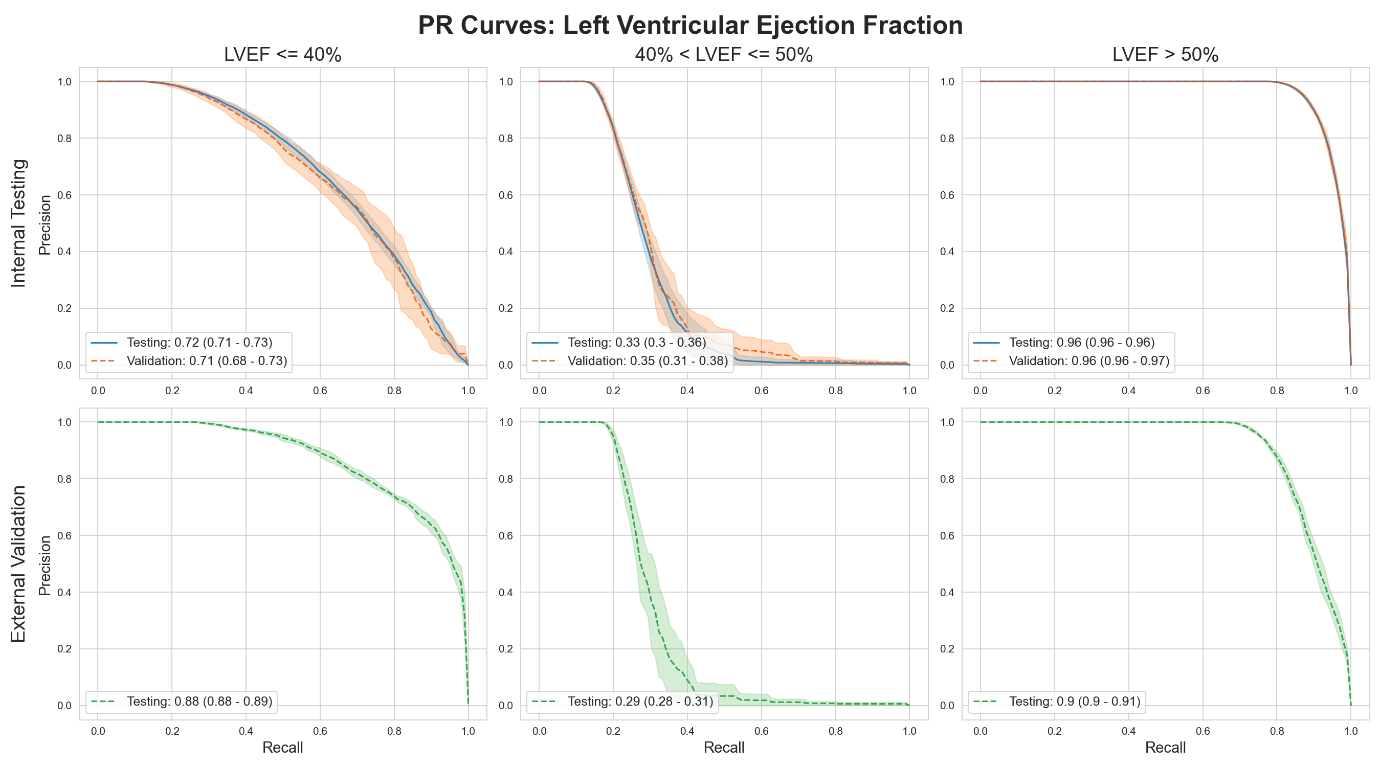

**Supplementary Figure 9**: Receiver Operating Characteristic curves for model performance with varying severity of Mitral Regurgitation.

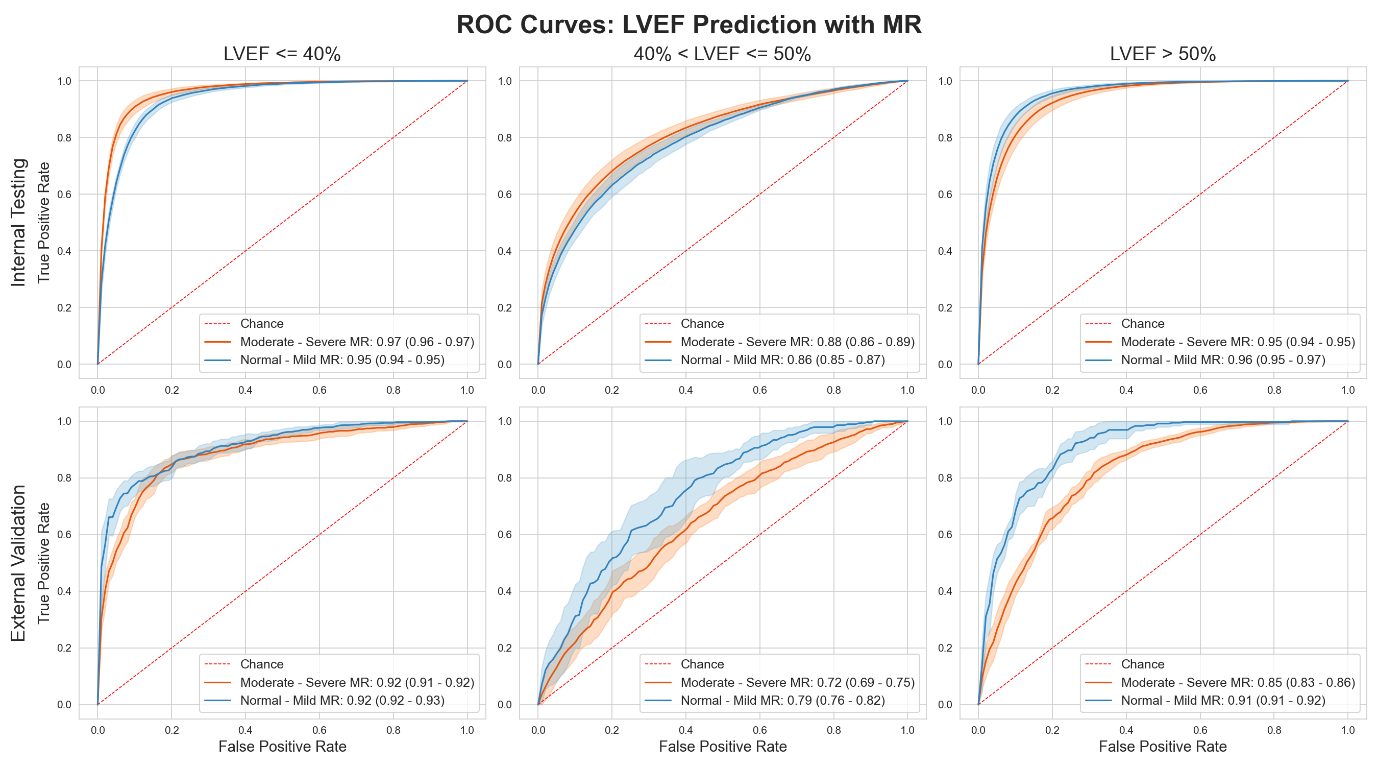

**Supplementary Figure 10**: Precision Recall curves for model performance with varying severity of Mitral Regurgitation.

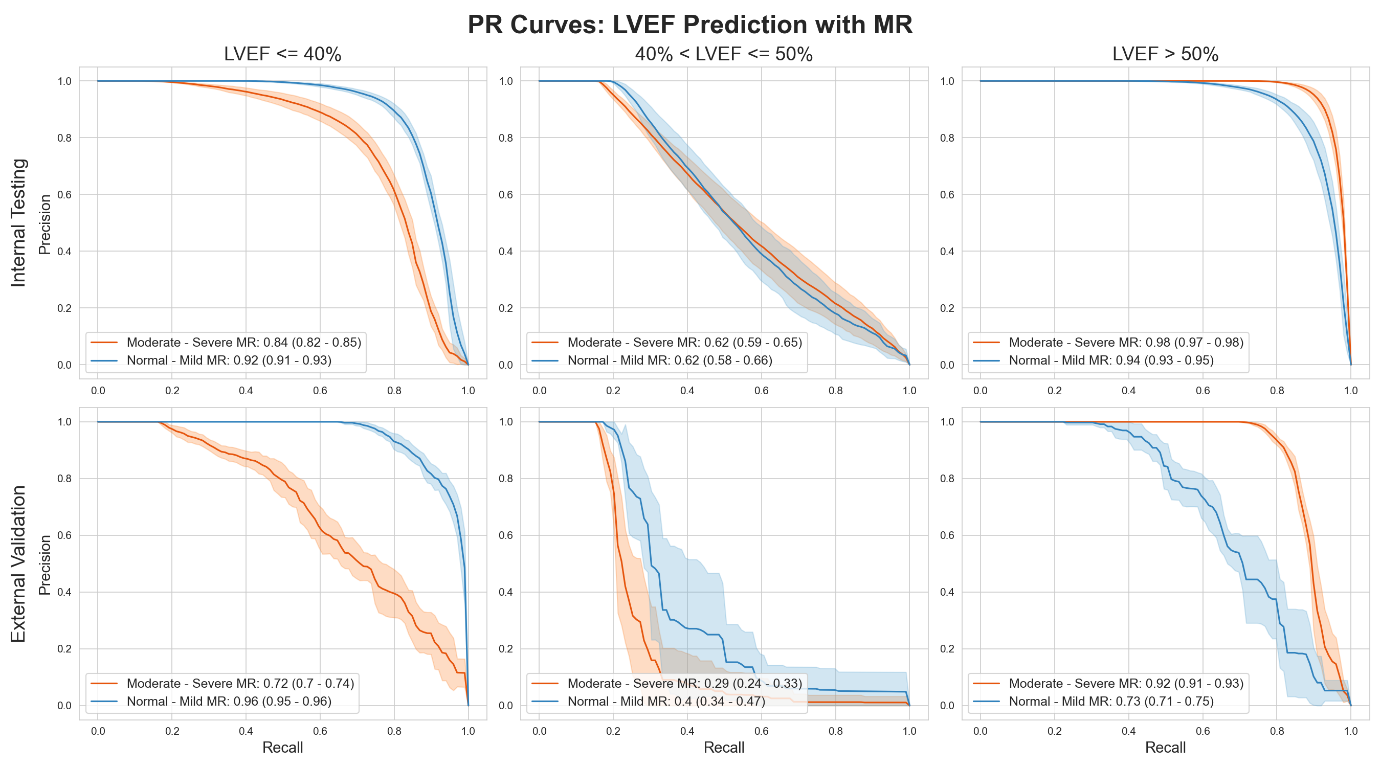

**Supplementary Figure 11:** Receiver Operating Characteristic curves for classification of LVEF <= 35%

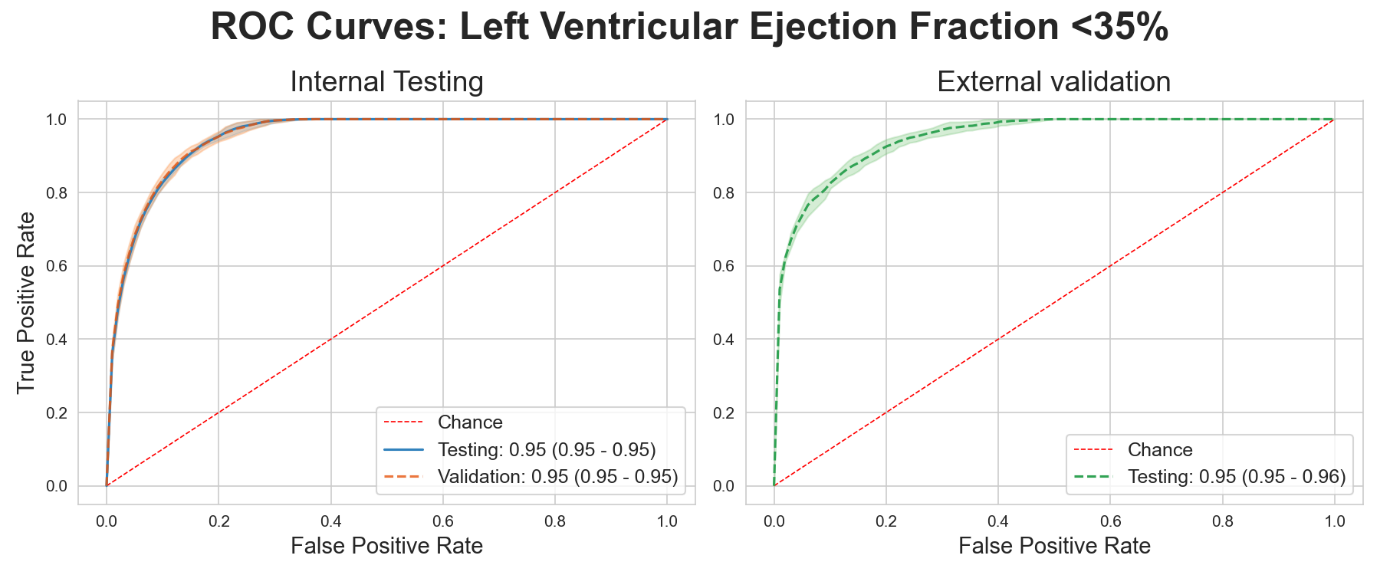

**Supplementary Figure 12:** Precision Recall curves for classification of LVEF <= 35%

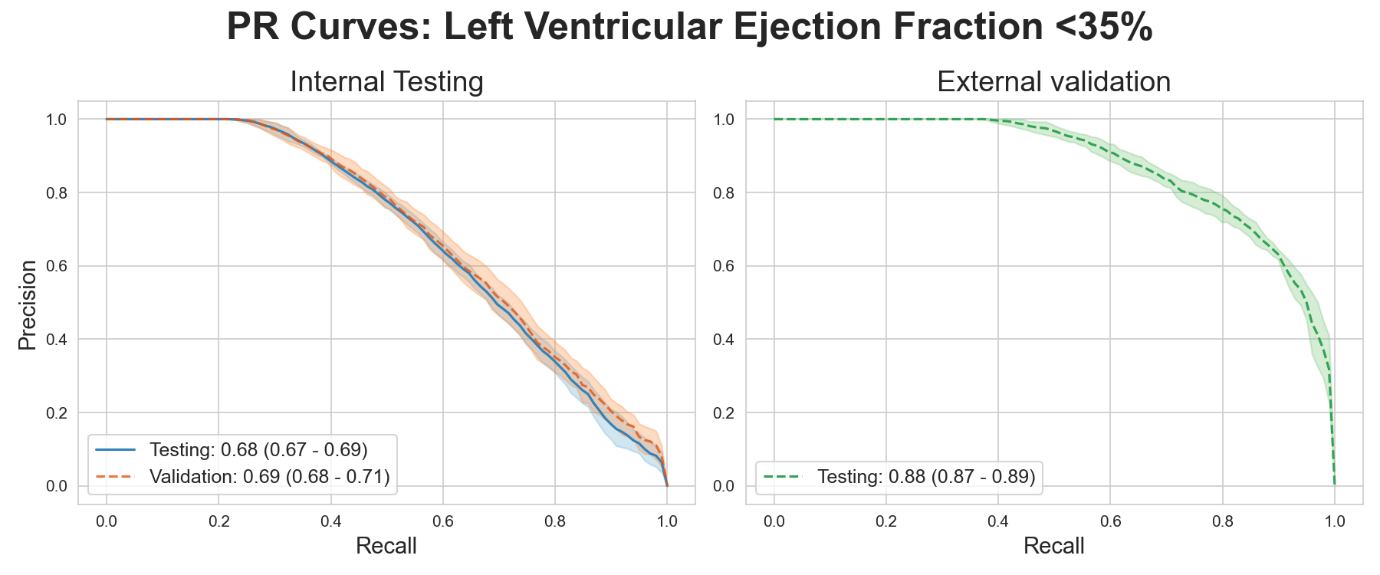

**Supplementary Figure 13:** Scatterplots showing LVEF regression performance in 3 subgroups of <40%, 40 - 50%, and >50%. Contour lines show density of predicted LVEF values vs ground truth LVEF values around the line of perfect concordance. Error bars around each datapoint indicate the mean absolute error. 2000 samples are shown for each panel to prevent overplotting. Contour maps were generated using the entire dataset.

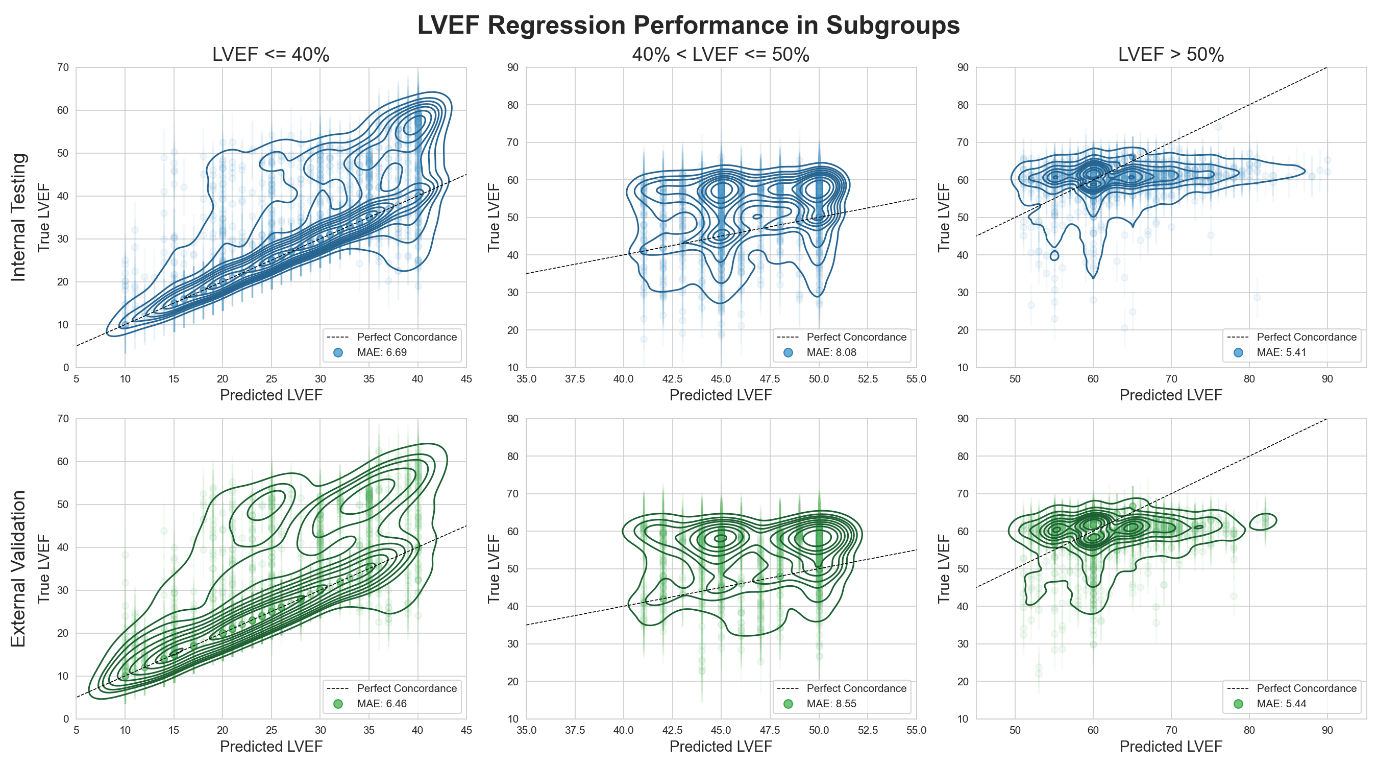

**Supplementary Figure 14**: Precision Recall Curves for RVD + RVSD classification

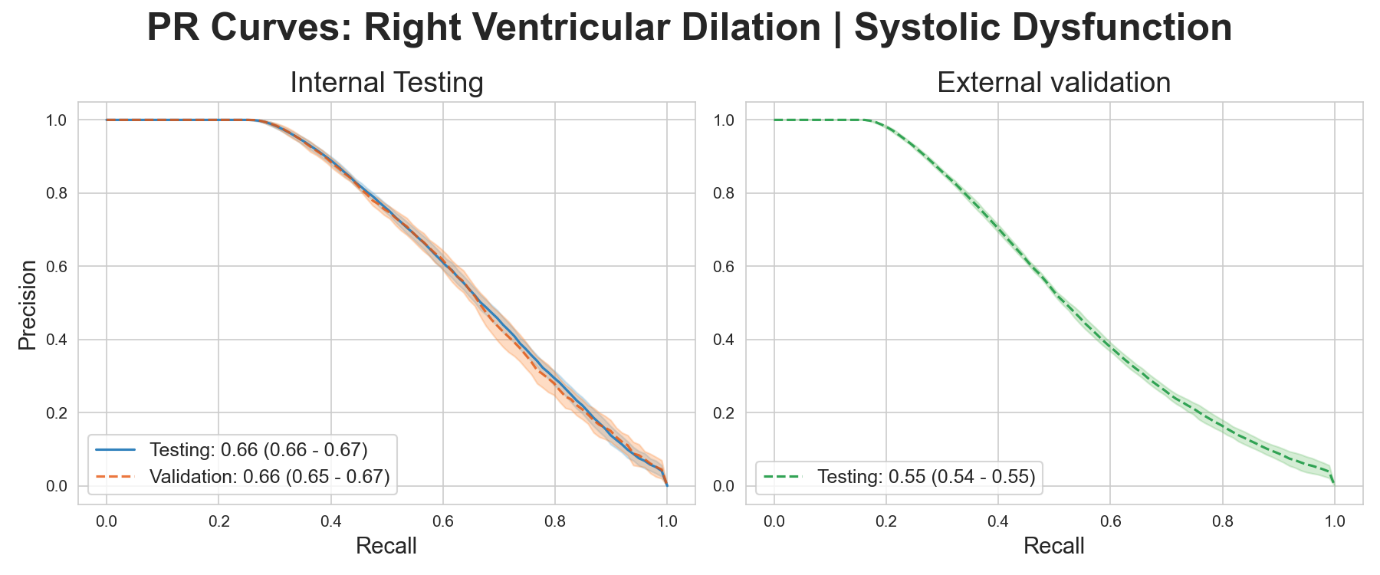

**Supplementary Figure 15**: Receiver Operating Characteristic curves for RVD + RVSD status prediction in presence of very low (<=40%) or close to normal LVEF (50%<).

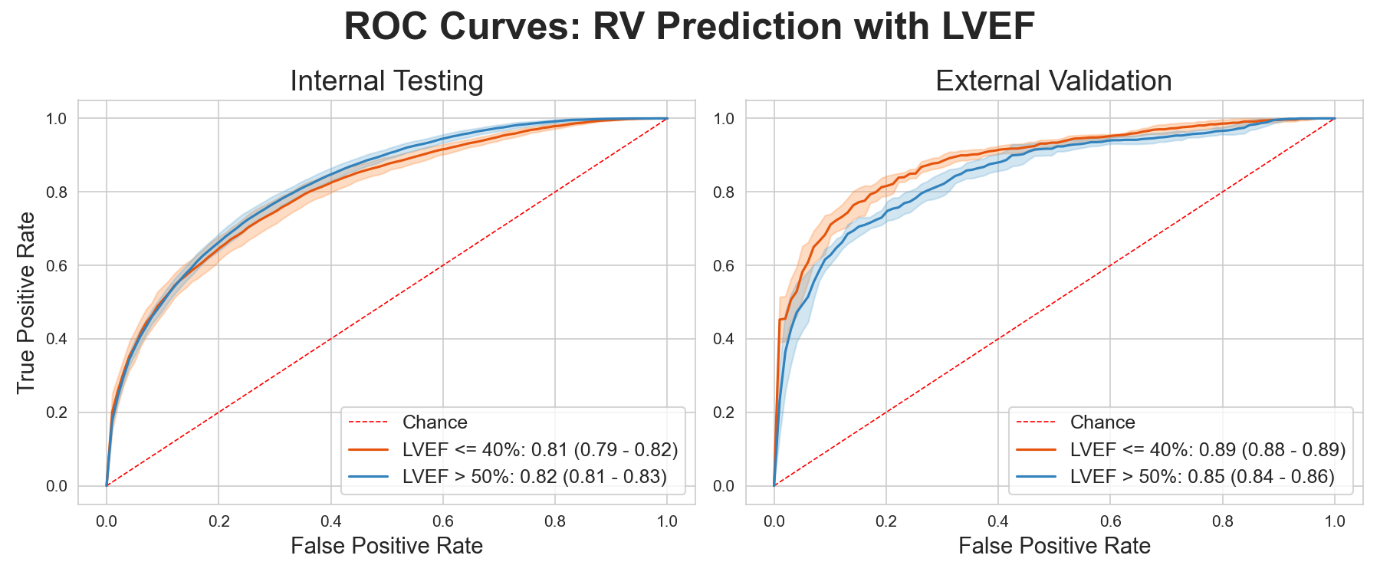

**Supplementary Figure 16**: Precision Recall curves for RVD + RVSD status prediction in presence of very low (<=40%) or close to normal LVEF (50%<)

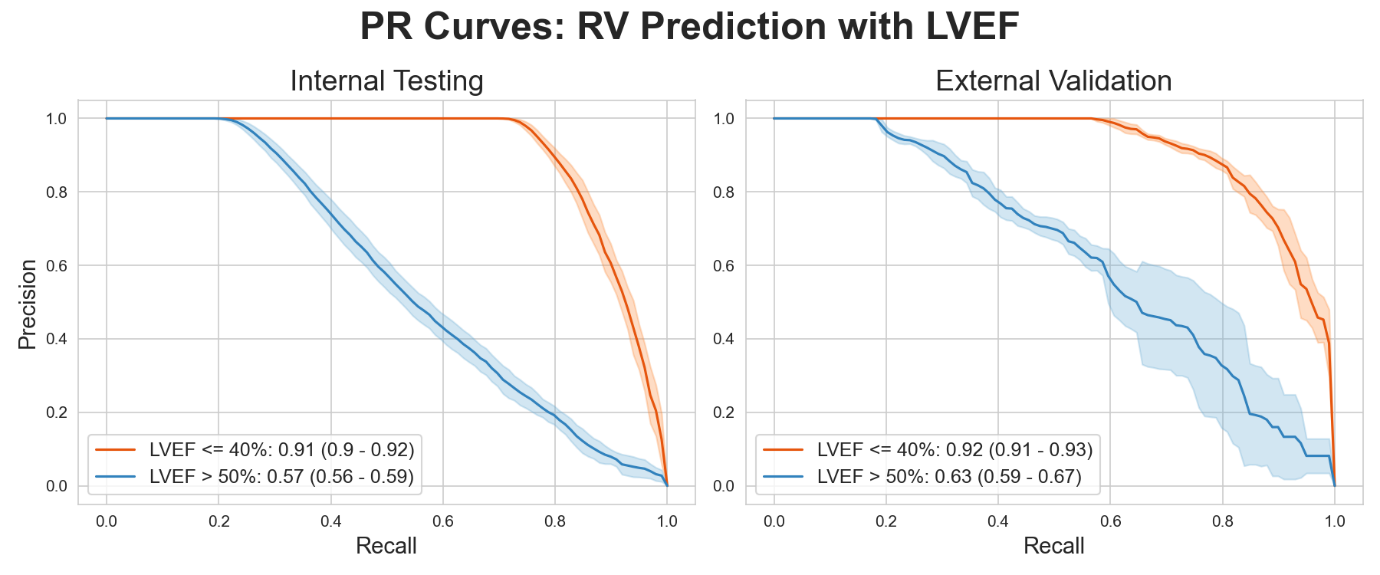

Supplementary Tables

**Supplementary Table 1:** NLP Rules

Natural Language Processing rules and sample positives for each rule. Terms in curly brackets imply special processing. *: wildcard, *LEMMA*: Modifications of word allowed e.g., reduce -> reduced. *|* : Either of these words. *STOP_WORD*: Words otherwise filtered out (such as *and*). *NEGATE*: Must not have the following word.

| **Rule** | **Example Positive** |
| --- | --- |
| **RV Normal Size** | |
| normal right {ventricle \| ventricular} {*} size | Normal right ventricular size |
| right {ventricle \| ventricular} {*} normal {*} {LEMMA:size} | Right ventricle is normal sized |
| rv size {*} normal | RV size is normal |
| right ventricle {*} {*} {*} {*} of normal {*} | Right ventricle appears to be of normal size |
| **RV Normal Function** | |
| normal right {ventricle \| ventricular} function | Normal right ventricular function |
| right {ventricle \| ventricular} wall motion {*} normal | Right ventricular wall motion is normal |
| normal right ventricular wall motion | Normal right ventricular wall motion |
| right ventricular function {*} normal | Right ventricular function is normal |
| **RV Size and Function normal** | |
| normal right {ventricle \| ventricular} size {STOP_WORD} {*} function | Normal right ventricular size and systolic function |
| right ventricle {*} {*} {*} {*} normal {*} size {STOP_WORD} function | Right ventricle is observed to have normal size and function |
| **RV Abnormal Size** | |
| dilated right ventricle | Dilated right ventricle |
| right ventricular {dilation \| dilatation} | Right ventricular dilation |
| rv size {*} {*} dilated | RV size is moderately dilated |
| right ventricle {STOP_WORD} {NEGATE: not} dilated | Right ventricle is very mildly dilated |
| **RV Abnormal Function** | |
| normal right {ventricle \| ventricular} size {STOP_WORD} {LEMMA:reduce} | Normal right ventricular size and reduced function |
| right {ventricle \| ventricular} {*} {LEMMA:reduce} systolic function | Right ventricle has reduced systolic function |
| {decreased \| reduced} right {ventricle \| ventricular} {*} function | Decreased right ventricular systolic function |
| right {ventricle \| ventricular} {*} function {*} {mildly \| moderately \| severely} | Right ventricular function is moderately reduced |
| right ventricle {*} {hypokinetic \| akinetic} | Right ventricle is hypokinetic |

**Supplementary Table 2**: NLP performance

Outcomes were selected equally from NLP detected *Normal / Abnormal / No mention in report*. The *Total* column includes outcomes missed by the NLP algorithm. Such reports and any matched ECGs were excluded.

| **Outcome** | **Normal** | | | **Abnormal** | | | **No mention in report** | | |
| --- | --- | --- | --- | --- | --- | --- | --- | --- | --- |
|  | ✓ | ✖ | Total | ✓ | ✖ | Total | ✓ | ✖ | Total |
| **Reviewer I** | | | | | | | | |  |
| RV Function | 123 | 0 | 127 | 44 | 0 | 45 | 36 | 2 | 38 |
| RV Size | 128 | 0 | 130 | 43 | 1 | 50 | 30 | 0 | 30 |
| Mitral Regurgitation | 23 | 0 | 23 | 149 | 2 | 157 | 28 | 2 | 30 |
| **Reviewer II** | | | | | | | | |  |
| RV Function | 120 | 0 | 125 | 49 | 1 | 52 | 32 | 0 | 33 |
| RV Size | 136 | 0 | 140 | 33 | 0 | 37 | 32 | 0 | 33 |
| Mitral Regurgitation | 17 | 0 | 21 | 148 | 3 | 162 | 27 | 0 | 27 |

**Supplementary Table 3:** LVEF classification performance in presence of moderate to severe mitral regurgitation OR normal to mild mitral regurgitation.

| **Outcome** | **Cohort** | **Mitral Regurgitation** | **% eval prevalence** | **AUROC** | **AUPRC** | **Sensitivity** | **Specificity** |
| --- | --- | --- | --- | --- | --- | --- | --- |
| LVEF <= 40% | Internal Testing | Mod./Severe | 12.57% | 0.97 (0.96 - 0.97) | 0.84 (0.82 - 0.85) | 0.92 (0.91 - 0.93) | 0.89 (0.88 - 0.91) |
|  |  | None/Mild | 39.43% | 0.95 (0.94 - 0.95) | 0.92 (0.91 - 0.93) | 0.91 (0.89 - 0.92) | 0.85 (0.84 - 0.87) |
|  | External Validation | Mod./Severe | 15.95% | 0.92 (0.91 - 0.92) | 0.72 (0.70 - 0.74) | 0.85 (0.82 - 0.88) | 0.84 (0.81 - 0.87) |
|  |  | None/Mild | 64.36% | 0.92 (0.92 - 0.93) | 0.96 (0.95 - 0.96) | 0.81 (0.78 - 0.84) | 0.91 (0.87 - 0.94) |
| LVEF 40 - 50% | Internal Testing | Mod./Severe | 16.17% | 0.88 (0.86 - 0.89) | 0.62 (0.59 - 0.65) | 0.80 (0.76 - 0.84) | 0.77 (0.74 - 0.79) |
|  |  | None/Mild | 20.45% | 0.86 (0.85 - 0.87) | 0.62 (0.58 - 0.66) | 0.81 (0.76 - 0.85) | 0.72 (0.67 - 0.76) |
|  | External Validation | Mod./Severe | 15.28% | 0.72 (0.69 - 0.75) | 0.29 (0.24 - 0.33) | 0.86 (0.78 - 0.94) | 0.5 (0.44 - 0.56) |
|  |  | None/Mild | 15.84% | 0.79 (0.76 - 0.82) | 0.4 (0.34 - 0.47) | 0.86 (0.77 - 0.94) | 0.64 (0.52 - 0.76) |
| LVEF >50% | Internal Testing | Mod./Severe | 71.28% | 0.95 (0.94 - 0.95) | 0.98 (0.97 - 0.98) | 0.89 (0.87 - 0.91) | 0.85 (0.84 - 0.87) |
|  |  | None/Mild | 40.33% | 0.96 (0.95 - 0.97) | 0.94 (0.93 - 0.95) | 0.91 (0.90 - 0.92) | 0.84 (0.82 - 0.85) |
|  | External Validation | Mod./Severe | 68.77% | 0.85 (0.83 - 0.86) | 0.92 (0.91 - 0.93) | 0.81 (0.75 - 0.86) | 0.73 (0.67 - 0.78) |
|  |  | None/Mild | 19.80% | 0.91 (0.91 - 0.92) | 0.73 (0.71 - 0.75) | 0.89 (0.84 - 0.94) | 0.82 (0.78 - 0.86) |

**Supplementary Table 4:** RV Systolic Dysfunction or RV Dilation performance in presence of LVEF <= 40% OR LVEF > 50%.

| **Cohort** | **LVEF Status** | **% eval prevalence** | **AUROC** | **AUPRC** | **Sensitivity** | **Specificity** |
| --- | --- | --- | --- | --- | --- | --- |
| Internal Testing | LVEF <= 40% | 70.93% | 0.81 (0.79 - 0.82) | 0.91 (0.9 - 0.92) | 0.71 (0.66 - 0.77) | 0.75 (0.7 - 0.8) |
|  | LVEF > 50% | 19.49% | 0.82 (0.81 - 0.83) | 0.57 (0.56 - 0.59) | 0.74 (0.72 - 0.76) | 0.73 (0.72 - 0.75) |
| External Validation | LVEF <= 40% | 55.90% | 0.89 (0.88 - 0.89) | 0.92 (0.91 - 0.93) | 0.8 (0.76 - 0.84) | 0.84 (0.8 - 0.89) |
|  | LVEF > 50% | 17.04% | 0.85 (0.84 - 0.86) | 0.63 (0.59 - 0.67) | 0.72 (0.69 - 0.76) | 0.85 (0.82 - 0.88) |
